# Influence of open-source virtual-reality based gaze training on navigation performance in Retinitis pigmentosa patients in a crossover randomized controlled trial

**DOI:** 10.1101/2023.09.11.23295342

**Authors:** Alexander Neugebauer, Alexandra Sipatchin, Katarina Stingl, Iliya Ivanov, Siegfried Wahl

## Abstract

**Motivation:** People living with tunnel vision caused by Retinitis pigmentosa (RP) oftentimes face challenges navigating and avoiding obstacles due to a severely decreased Visual Field. Previous studies have shown that gaze training – guided tasks to encourage specific gaze behavior – help improve visual performance in patients with limited Visual Field. We propose a Virtual-Reality based, at-home gaze training – to the best of our knowledge the first of its kind – to increase effective eye movements and navigation performance of RP patients in a real-world obstacle course.

**Methods:** A group of RP patients (n=8) participated in a study consisting of two 4-week-phases, both carried out by the same patient group in randomized order: In the ‘training phase’, participants carried out a Virtual-Reality gaze training for 30 minutes per day; In the ‘control phase’, no training occurred. Before and after each phase, participants were tasked to move through a randomized real-world obstacle course. Navigation performance in the obstacle course as well as eye-tracking data during the trials were evaluated. The study is registered at the German Clinical Trials Register (DRKS) with the ID DRKS00032628.

**Results:** On average, the time required to move through the obstacle course decreased by 17.0% after the training phase (*p <* 0.001), the number of collisions decreased by 50.0% (*p <* 0.001) and the average visual area observed by participants increased by 4.41% (*p <* 0.001). In comparison, after the control phase, the time required to move through the obstacle course was found to have decreased by 5.9% (*p* = 0.003), collisions decreased by 10.4% (*p* = 0.609) and the observed visual area increased by 2.06% (*p* = 0.0835).

**Conclusion:** The performance increase over the training phase significantly surpasses the natural learning effect found in the control phase, suggesting that Virtual-Reality based gaze training can improve real-world navigation performance and effective gaze movements of patients with RP. The training is available as work-in-progress open-source software.

## Introduction

Retinitis pigmentosa (RP) is a subset of inherited retinal diseases characterized by progressive loss of the Visual Field (VF) due to the degeneration of the retina [1–3]. This loss of the VF starts at the periphery or middle periphery and leads to blindness in the long-term progression of the degeneration [4, 5]. Other symptoms of RP include blurriness of sight, glare sensitivity, as well as night blindness [1, 6]. RP is estimated to occur in about 1 in 4000 people [5, 7–9].

The condition in which visual information can only be perceived in the center of the VF is known as “tunnel vision”. It can have severe impact on the daily lives of those affected by RP [6], especially in visual tasks such as navigation and visual search. At the time of writing there is only one approved gene therapy for retinitis pigmentosa [10]. Despite good results in efficacy of this therapy also on the visual field, in the majority of the patients halting the progression is not consistently possible [6, 8]. It is therefore essential to explore other methods that can improve the visual capabilities of RP patients - and thus improve their quality of life.

One of these approaches is gaze training, which involves teaching patients how to adjust their gaze movements to compensate for their missing visual areas [11, 12]. For patients with limited VF, an important technique for this approach is the use of exploratory saccades. Exploratory saccades are rapid eye movements that help explore the visual environment by quickly shifting the point of fixation to new locations [13]. While these eye movements can not directly improve the biological health of the retina or increase the size of the ”static” VF, i.e. the visual area that can be perceived at any one time, they can increase the visual area that is observed over time, facilitating the detection of new visual information. By incorporating exploratory saccades into gaze training, patients can learn to adapt their gaze movements to partly accommodate for their limited VF and observe larger areas around them. This can lead to better obstacle detection, safer navigation, and an overall higher level of independence in everyday visual tasks.

The concept of gaze training for low-vision compensation has been investigated and applied before [11–15]. However, the constant advancements in technology and accessibility of Virtual Reality (VR) headsets raises a question about their potential to be applied for gaze training purposes. Compared to conventional, computer display based setups, VR devices offer a number of possible advantages.

- The displays of a VR headset cover larger visual angles than a traditional computer screen, with most commercially available VR headsets featuring visual angles of 90° per eye or higher [16]. Assuming the recommended minimal distance from a working screen of 50cm [17], a 45” screen (99.7cm×56.0cm) is required to match the visual angle of a VR headset at least in horizontal dimension, and an 80” computer screen (177cm×99.6cm) would be required to also match the vertical visual angle.
- In addition, VR headsets can measure head rotations and adjust the displayed image in real-time to mimic the effect of “looking around”. This further increases the visual angles at which VR headsets can display visual content, allowing for a full 360° view.
- Lastly, the use of VR allows the risk-free simulation of immersive, interactive 3D environments that provide a perspective and visual experience much closer to that of the real world.

To the best of our knowledge, at the point of writing there is no Virtual Reality based gaze training for people with visual field deficit apart from the one presented in this work. However, research on the use of Virtual Reality for adjusting gaze behavior in other fields, such as for industry task training [18] or as therapeutical intervention for patients with mental health disorders [19], suggests that the use of VR applications is feasible to influence gaze behavior. In addition, it has been shown that skills trained in VR can have sustained transfer effect to real-world performances [20–23]. In this work, we are investigating the potential of Virtual Reality to be applied for unsupervised, at-home gaze training, as well as the influence of gaze training in a virtual environment on the navigation performance in real-world tasks.

## Materials and methods

The first part of this section will focus on the developed gaze training, its implementation and the intentions behind its design. Subsequently, an experimental study will be presented to show how the gaze training impacted real-world navigation. A CONSORT flowchart for this study is provided in Fig 1.

**Fig 1.**
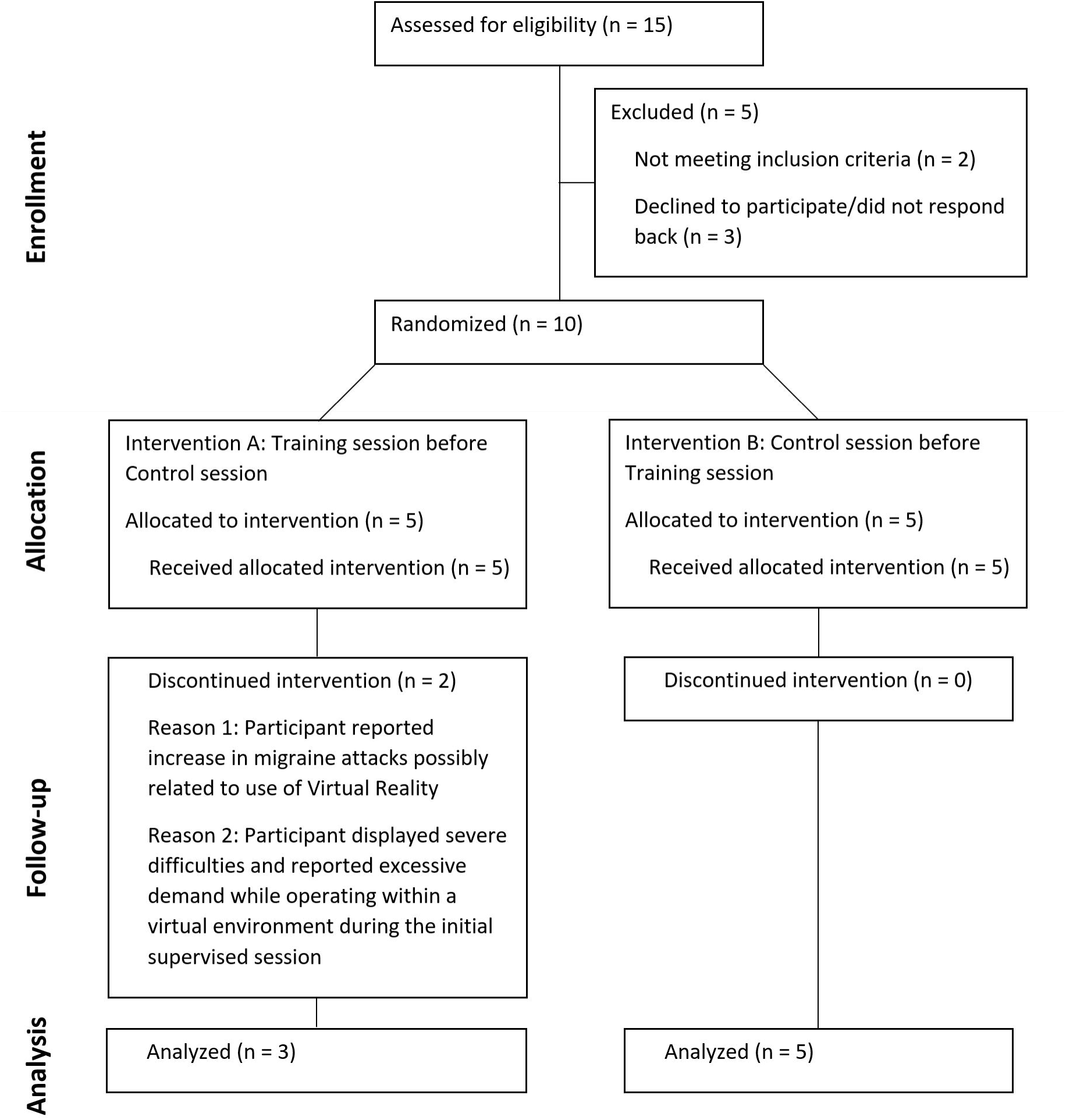
CONSORT flowchart. The CONSORT flowchart for the patient study described in this work.

### Development of a virtual-reality based gaze training tool

The initial phase of our project was dedicated to the development of the gaze training software. The aim was to create a tool that is easy to use, engaging, and that provides visual training tasks to motivate larger and more frequent eye movements. The training should be usable in an unsupervised at-home environment. Thus, it had to be ensured that no physical movement is required that would put the user or their environment at risk. All tasks were designed to be executed in seated or stationary standing position: While the viewing direction within the VR environment was controlled physically through head and body rotation, any form of locomotion was triggered solely through controller input.

#### Software and hardware specifications

The training software was developed in the Unity3D game engine (Version 2021.3LTS), using the Pico XR SDK (version 1.2.4). The Pico Neo 2 Eye VR headset was used for development and training. It provides stand-alone functionality, meaning that no connection to a computer or any external tracking devices is necessary. The device features a 75 Hz display refresh rate and the VF per eye is stated to be 101° [24] according to the developer’s specifications, though independent measurements have shown a VF per eye of 89° both horizontally and vertically [16]. The built-in eye tracker of the Pico Neo 2 Eye has a refresh rate of 90 Hz and an accuracy of 0.5° according to the device specifications, with an ideal eye-tracking range of 25° horizontally and 20° vertically [24]. The use of VR is possible while wearing glasses or contact lenses.

#### Training tasks

The training software consists of three visual tasks, each one designed to promote exploratory saccades and frequent eye movements:

- **Target tracking** In this task, a varying number of targets (starting at five) move across a two-dimensional area in front of the user in a random pattern (Fig 2 A and D). To make the task more visually appealing and thematic, targets were displayed as cartoon-styled mice. At the start of training, the area’s dimensions are 52° horizontally and 39° vertically, which roughly represents 30% of a healthy VF at 180°×135°. Two of the targets are marked at the start of the trial (visualized as a piece of cheese carried by the mouse, as illustrated in Fig 2 A), and the user is asked to follow the marked targets with their gaze in order to not lose track of them. After 8-12 seconds, all targets stop their movements, and the marked targets change their appearance to become indistinguishable from the non-marked targets. At this point, the user is prompted to select the two formerly marked targets through input of the VR controller. Selected targets are revealed to be either correct or incorrect. A trial is considered to be successful if the user selects both correct targets and no or only one incorrect target. When selecting two incorrect targets, the trial fails.
- **Search Task** In this task, participants are given a 20-second time limit to search an area in front of them. As in the Target Tracking Task, the default dimensions of this area are 52° horizontally and 39° vertically. The objective of the task is to search for stationary targets, marked by a prominent cross symbol (Fig 2 B and E), and to use the controllers to select as many of them as possible within the given time frame. A total of three marked targets are placed in the defined area and once a target is selected, it is instantly moved to a new position inside the designated area, with a minimum distance of 30° from the previous position. This minimum distance is introduced to avoid targets re-appearing directly in the participant’s VF, thus further promoting continuous scanning of the area to find additional targets. In addition to the targets marked with a cross, there are similar targets marked with a circle that serve as distractors to ensure that participants fully focus on a target before selecting it. Each trial was rated based on the number of marked targets that are found within the limited time frame.
- **Navigation Task** In the third task, participants are asked to navigate through a randomized obstacle course simulated in a virtual environment (Fig 2 C and F). Using the controllers of the VR device, the participant is able to move at a dynamic pace, with a set maximum speed of 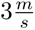 by default. The movement direction is controlled via the participant’s body orientation, which is measured through the VR headset’s orientation. The obstacle course is designed as a corridor with two left turn tiles, two right turn tiles and two straight tiles, each measuring 8 meters in both width and length (Fig 3). The six tiles are arranged in randomized order for a total of 90 unique layouts. Along the corridor, 12 randomized obstacles are placed. To motivate adaptive eye movements, obstacles have different height, shape and movement patterns and can be divided into three categories: Near-ground obstacles require scanning of the lower visual area; Obstacles hanging at head-level require eye movements towards the upper regions of the visual area; Moving obstacles periodically move from one side of the walking corridor to the other and thus require dynamic and frequent eye movements to notice and avoid them. Collisions with obstacles or the walls bordering the walking corridor are indicated through a sound cue as well as a “bouncing” animation that moves the user’s avatar back slightly. The goal of the obstacle course is marked with a prominent red circle, which the participant has to reach in order to successfully finish the trial. The trial was rated based on the duration required to move through the obstacle course as well as the number of collisions during the trial.

**Fig 2.**
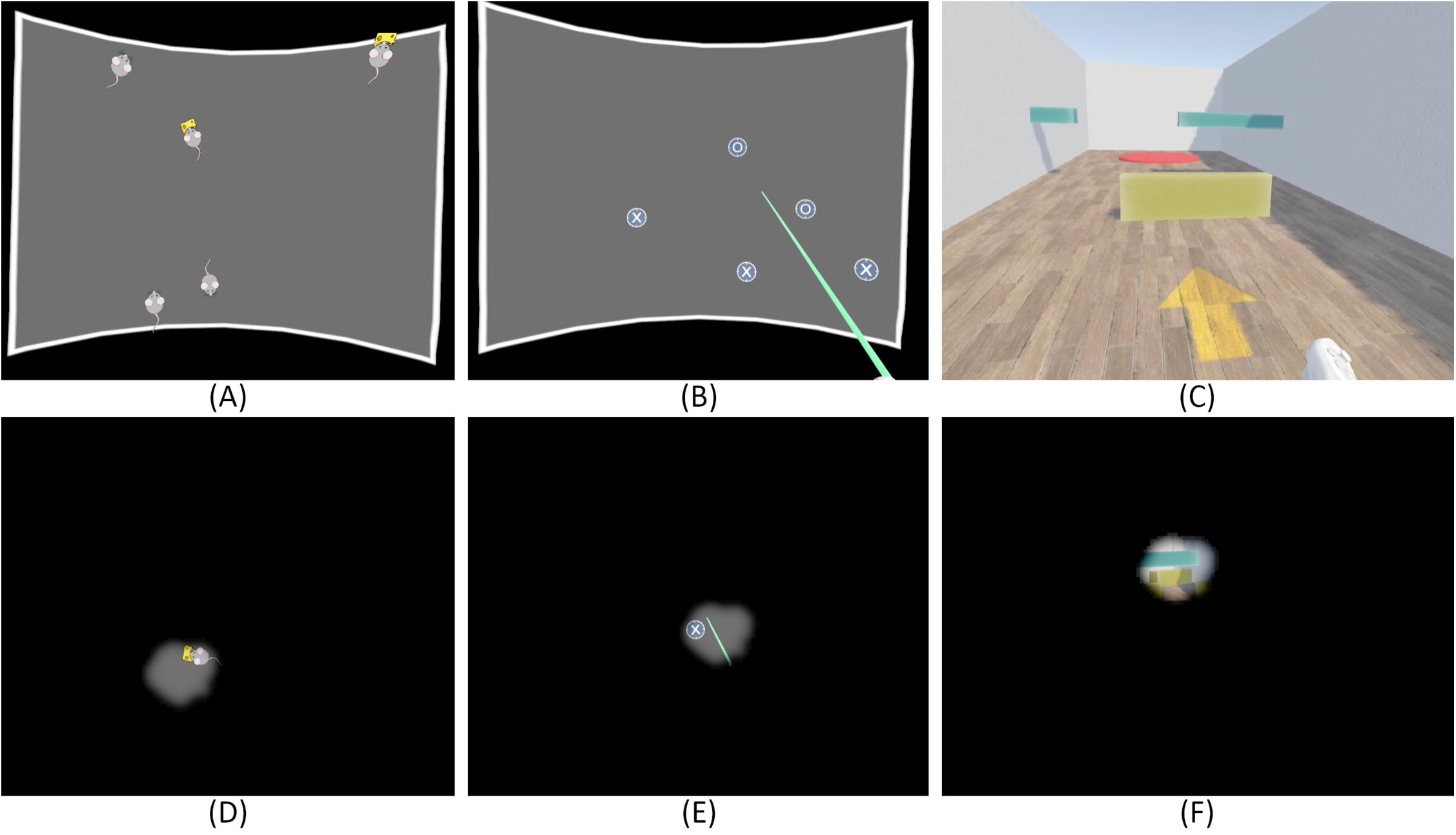
Screen captures of the three visual tasks of the VR gaze training. A: Target Tracking (marked targets are indicated by a piece of cheese); B: Search Task; C: Navigation Task; D: Target Tracking with simulated tunnel vision; E: Search Task with simulated tunnel vision; F: Navigation Task with simulated tunnel vision. The tunnel vision displayed in D-F has a 15° diameter. The tunnel vision simulation is added for visualization purposes only and was not present during participants’ training.

**Fig 3.**
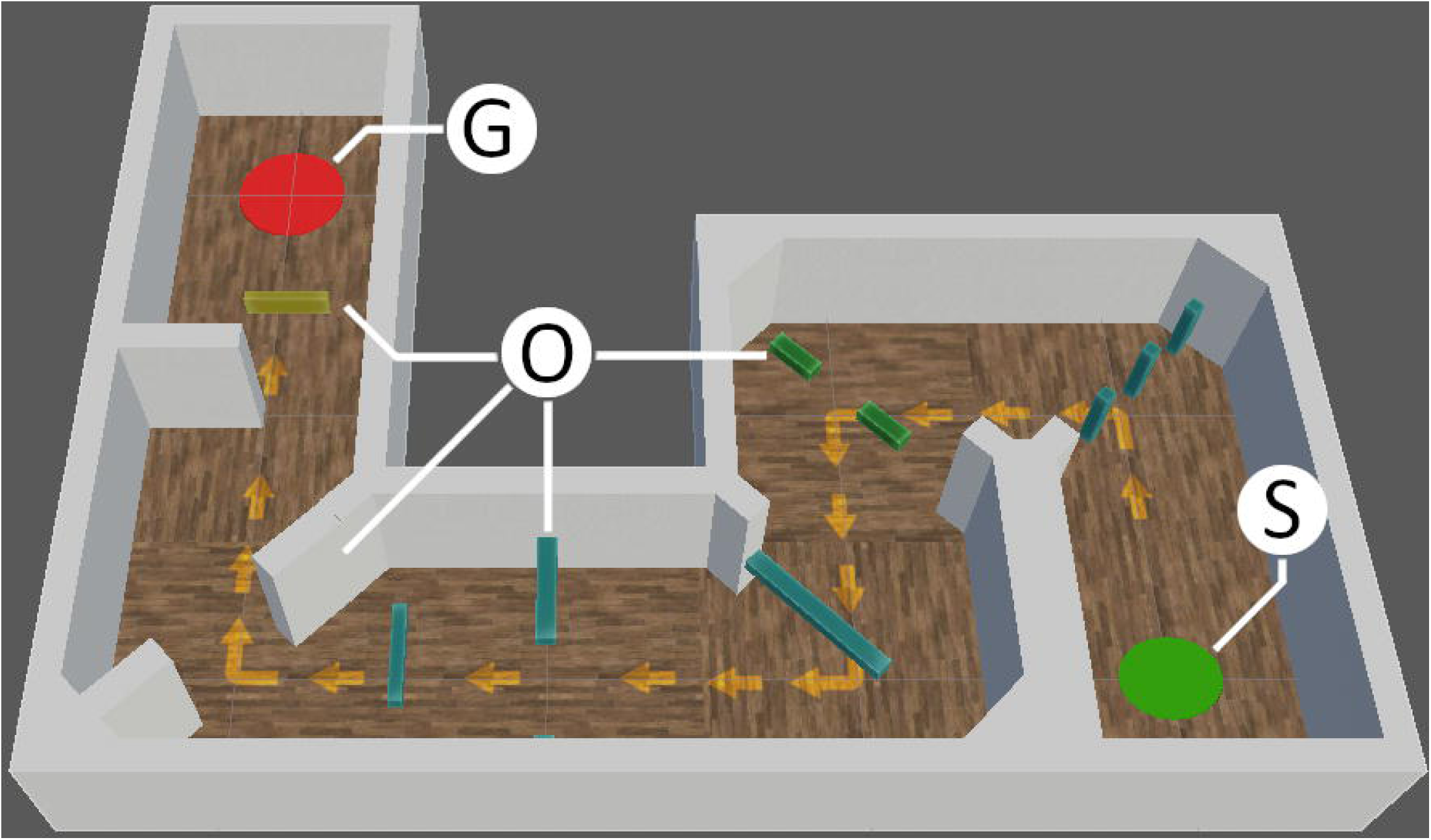
Navigation task visualisation. Top-down view on a randomized obstacle course of the Navigation Task. S: Starting position; O: Obstacles (example selection); G: Goal area.

After each trial, participants are brought back to a selection menu in which they are able to inspect the rating and result of the trial as well as their overall progress, go back to the main menu or start the next trial. Additionally, participants had the option to mark the previous trial as ”invalid”, but were instructed to only mark trials as invalid if there were technical or external factors distorting the results. A video showcasing the three training tasks can be found in the supporting files (S1 Video).

#### Adaptive difficulty levels

One of the design goals for the gaze training was to keep users engaged and motivated even throughout extended training phases. At the same time, the visual tasks should have low entry levels to make it easy for participants to get started and get used to the training tasks. To follow both premises, adaptive difficulty levels were introduced in all three visual training tasks. This means that the difficulty level of each individual task increases or decreases automatically based on the participant’s current performance in that task, with the aim to ensure that the tasks remain at an appropriate level of difficulty to keep the user engaged and motivated. In the selection menu in-between trials, participants are informed about the difficulty level they have reached and about their progress towards the next difficulty level.

For the Target Tracking Task, higher difficulty levels translate to a larger bounding area in which the targets move, higher target movement speed, and a greater number of both marked and unmarked targets. Similarly, increased difficulty in the Search Task leads to an expansion of the area in which targets are located, and increases the number of distractor targets while keeping the number of correct targets at three. In the Navigation Task, the maximum movement speed of the participant’s avatar gradually increases with higher difficulty level, while at the same time reducing the time limit to move through the obstacle course without reducing the performance rating.

Additionally, the speed of moving obstacles in the Navigation Task is adjusted to match the participant’s increased movement speed, resulting in a faster-paced trial that demands quicker reaction times and heightened situational awareness to avoid obstacles.

#### Scanpath behavior

Preliminary studies [13, 25] suggest the potential of specific systematic eye movements - called Scanpaths - to positively impact gaze training. Following this, participants were encouraged to follow a suggested, systematic Scanpath (visualized in Fig 4) while executing the training.

**Fig 4.**
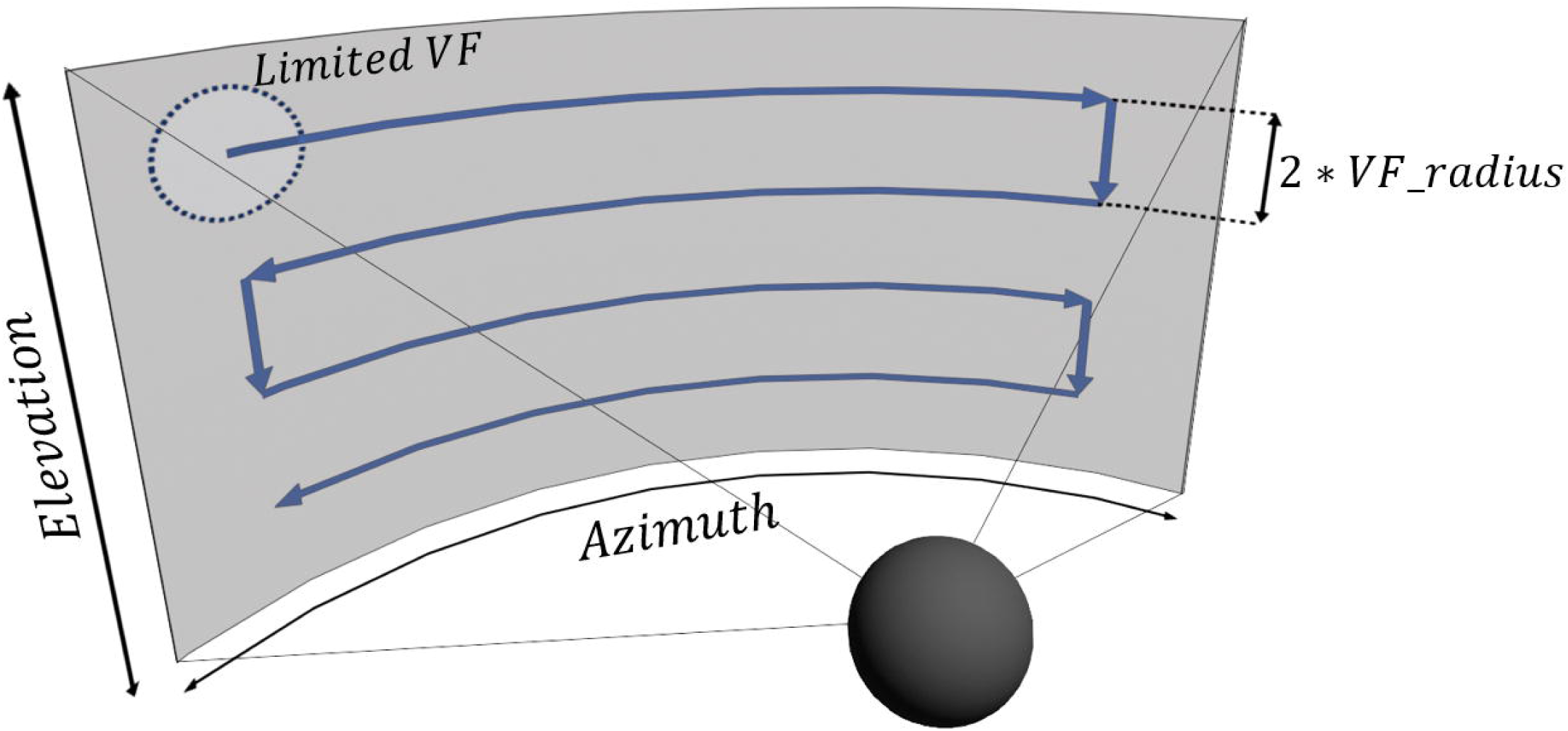
Scanpath visualization. Visualization of the pattern presented to the participants. The pattern starts in the upper left (or right) corner, moving along the azimuth axis to the opposite side. Then the pattern moves down along the elevation axis with an angular distance equal to the diameter of the participant’s VF. This pattern is continued until the lowest area was scanned. Following this pattern ensures that a large visual area is covered by the limited VF in an efficient manner.

Participants were given autonomy in choosing if - and to which degree - they want to follow the suggested Scanpath. It was presented on a screen and explained to them prior to the use of the gaze training. In addition, after each training trial within the VR environment, participants received automated feedback in form of a ”similarity value”, a quantitative measurement of how closely their real gaze behavior matched the suggested Scanpath. This quantitative measure of similarity between gaze behavior and suggested Scanpath is evaluated at run-time using a Multimatch algorithm [26], and is described in detail in Appendix A (S1 Appendix). The process behind choosing the pattern for the Scanpath, as well as the decision to make the execution of the Scanpath a voluntary option rather than a necessary part of training, is described in the discussion.

### Experimental study

To test the influence of gaze training on the navigation performance in the real world, we designed an experimental randomized controlled crossover study. The layout consisted of two phases (Fig 5):

- In the training phase, participants used the developed VR gaze training software at home for 10 hours over the course of 3-4 weeks (20 training sessions, 30 minutes per day, 10 minutes per task).
- In the control phase, participants would follow their normal life routine over a similar duration without carrying out any gaze training.

**Fig 5.**
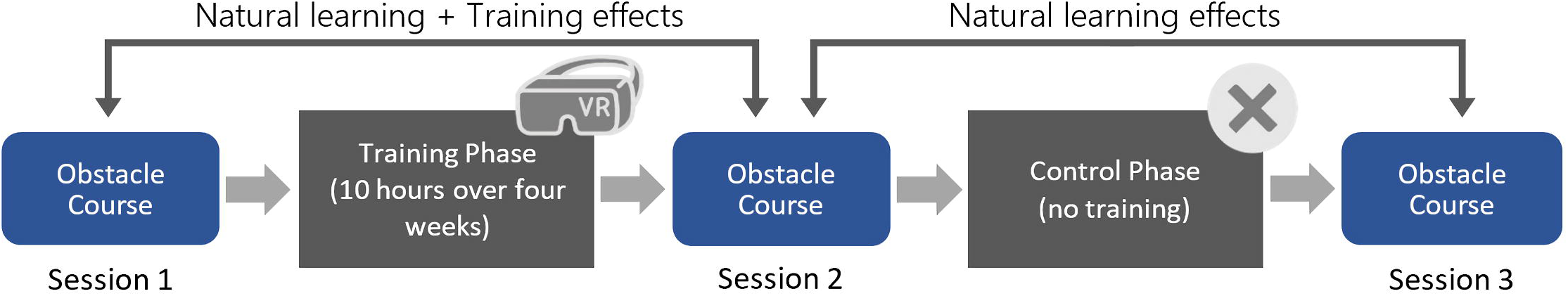
Study structure. Schematic of the study structure, including two phases of 3-4 weeks and the three in-person obstacle course sessions held. The order of training phase and control phase was randomized for each participant.

The experimenter randomized the order of the two phases for each participant during scheduling using a random number generator, allocating five participants to the group starting with training and five participants to the group starting with the control phase. Before and after each phase, participants completed an in-person session in which their task was to move through a randomized real-world obstacle course (20 trials per session). Details about the setup and experimental environment for these sessions and the obstacle course trials can be found in Experimental setup. The purpose of the control phase was to account for any improvements that might occur during the task that are not correlated to the gaze training. Despite randomizing the obstacle course setup to minimize memorization effects, participants can still be expected to learn and improve their performance by becoming familiar with the base structure and types of obstacles in the obstacle course. This natural improvement in performance is considered the ’natural learning effect’ of the experiment.

The impact of the training can thus be evaluated in three steps:

1. We assess the navigation performance in the real-world obstacle course before and after the training phase to determine the combined effect of both the potential training effect and natural learning effect. At this step, it is not yet possible to distinguish between the two effects.
2. Next, we assess the navigation performance before and after the control phase to find the natural learning effect displayed by participants, with no influence of gaze training.
3. By determining the differences between the “distilled” natural learning effect found in step 2 and the combined effect of training and natural learning found in step 1, it is possible to evaluate the effect that the developed gaze training has on real-world navigation performance. If the effect found after the training phase is significantly higher than the effect after the control phase, the gaze training can be considered successful.

#### Ethics and clinical trial registry

This study was proposed to and approved by the ethics committee of the Institutional Review Board of the Medical Faculty of the University of Tübingen (628/2018BO2) in accordance with the 2013 Helsinki Declaration. Patients were recruited in the time from June 9, 2022 to November 22, 2022. All participants signed informed consent forms. The study is registered as a clinical trial at the German Clinical Trials Register (DRKS) with the registry ID DRKS00032628. The registration was done retrospectively, as the study was originally considered as non-interventional observation study, not as clinical trial. Prompted by later feedback, this decision was reconsidered and the study was registered. The authors confirm that all ongoing and related trials for this intervention are registered.

#### Study population

10 patients (one male, nine female), with age ranges between 18-25 and 56-60 years (average 49.6 ± 13.0), participated in the study, two of which discontinued the study early on. Participation criteria were the diagnosis with Retinitis pigmentosa, a VF between 5° and 30° diameter, a visual acuity of 0.1 or higher and unrestricted mobility. It was tested in the first session that the participants are able to effortlessly see and recognize all targets and other visual features used in the gaze training, as well as all interfaces and menus required to operate the VR headset. The sample size was determined assuming a standard deviation of 20% of the mean navigation performance and an increase in performance of 25% after training, with an alpha of 0.05 and a power of 0.8.

Tab 1 lists information about the eight participants that finished the study. The provided medical data is based on the most recent medical examination of each patient. The medical examinations only provided visual representations of the VF of patients, which are included in Appendix B (S1 Appendix). In addition to these, the VF was measured during the first in-person session using a VR based kinetic perimetry developed for this project. While these measurements do not have diagnostic validity, they provide a better estimation of the perceived visual area of patients within the VR setup. This approach also ensured consistency in the measurement of VFs between participants.

#### Experimental setup

The real-world obstacle course was set up in an area with the dimensions of 4.8 meters width and 9.0 meters length. Two static privacy screens (visible in Fig 6) were placed such that an S-shaped corridor is formed. This extended corridor had a length of 18 meters – assuming a pathway exactly along the middle of the corridor – and a width of 3 meters.

**Fig 6.**
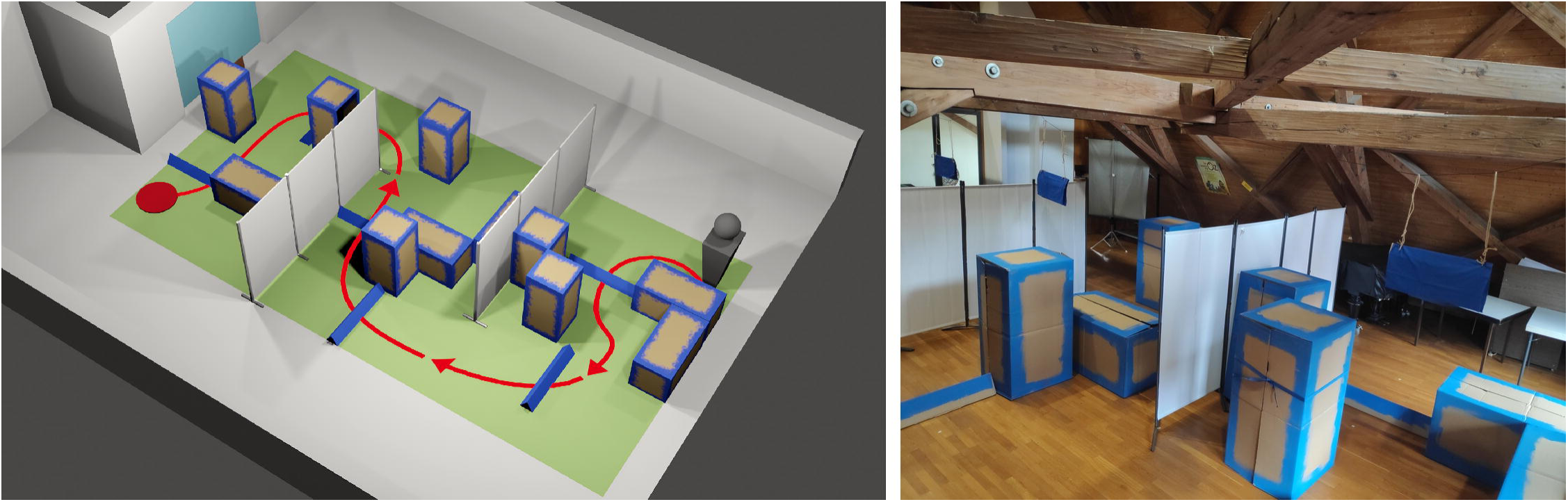
Obstacle course layout. Example of the real-world obstacle course in simulation (left) and actual setup (right). The simulated obstacle course is for presentation purposes only and was not used as part of the study.

Within the path, different obstacles were placed in a semi-randomized layout. Each obstacle arrangement consisted of 12 large carton boxes measuring 120×60×60 centimeters. Six of the boxes were oriented horizontally, six vertically. The set of obstacles also included six low-height obstacles that required participants to step over them, measuring 120×20 centimeters. Lastly, three sheets of cloth of 60 centimeters width were hanging from the ceiling, their lower edge at a height of 150 to 170 centimeters, adjusted to be on participants’ eye level. A total of 20 randomized obstacle layouts were created, with each layout being used exactly once per session. An example for one of these layouts is found in Fig 7. All obstacles were colored in blue to increase the contrast against the floor and background. The primary walking direction of the obstacle course was chosen such that participants were always facing away from the windows to avoid glare effects. The room was fully lit during all trials. A video showcasing the obstacle course in an example trial is found in the supporting files (S2 Video).

**Fig 7.**
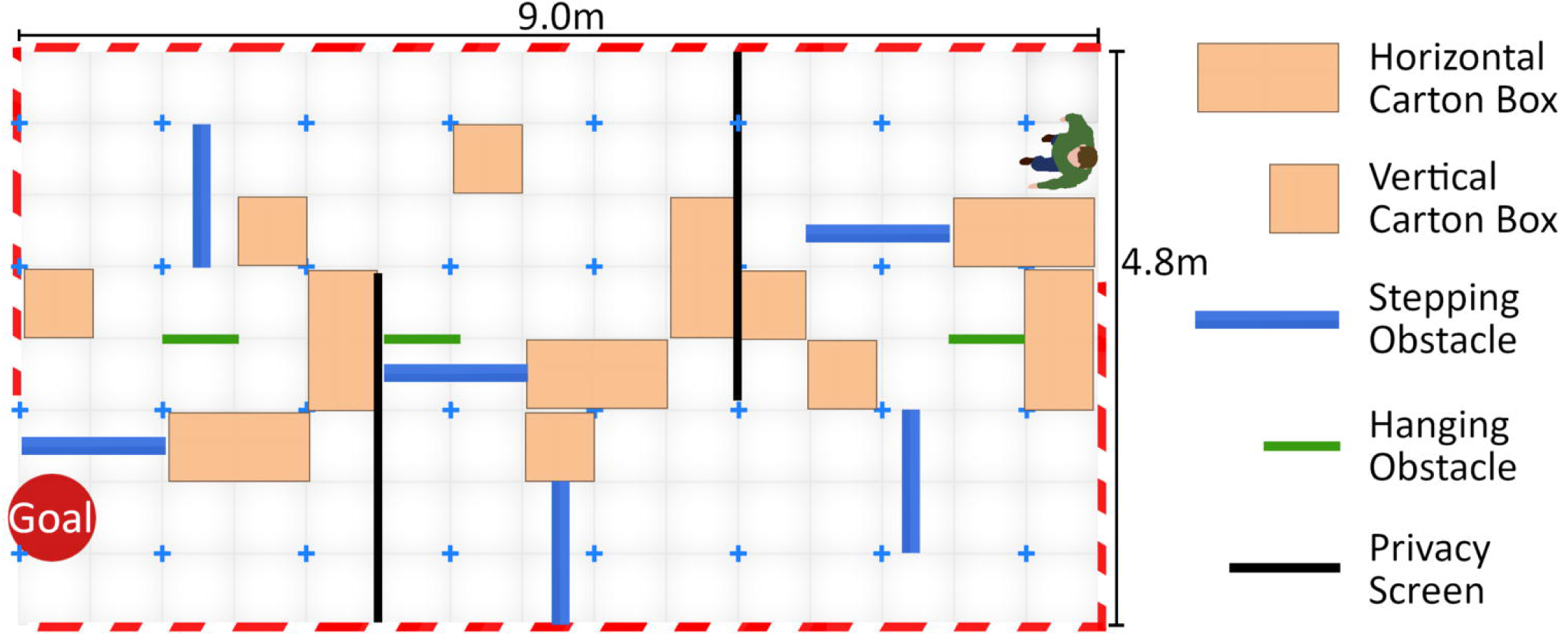
Obstacle course schematic. Example of one of the schematic layouts used to set up the obstacle course before each trial. Participants started each trial at the position that is marked by a person in the layout.

**Table 1.** Patient data.

| Patient<br>(Age / Sex) | Age of<br>diagnose | Visual field<br>(RE / LE) | Visual acuity<br>(RE / LE) | VF<br>notes | Gaze training<br>experience | VR<br>experience | Vision<br>correction |
| --- | --- | --- | --- | --- | --- | --- | --- |
| 1 (18-20f) | 11-15 | 7.62° / 8.26° | 0.40 / 0.40 | - | - | high | G/C<sup>1</sup> |
| 2 (56-60f) | 26-30 | 18.64° / 18.18° | 0.20 / 0.05 | spots<sup>2</sup> | VisioCoach | - | G |
| 3 (50-55f) | 16-20 | 17.64° / 16.36° | 0.13 / 0.20 | - | - | - | G |
| 4 (46-50m) | 21-25 | 24.60° / 25.40° | 0.05 / 0.05<sup>3</sup> | - | - | low | G |
| 5 (56-60f) | 46-50 | 18.54° / 18.34° | 0.32 / 0.25 | spots<sup>2</sup> | VisioCoach | - | G |
| 6 (56-60f) | 16-20 | 10.92° / 9.64° | 0.10 / 0.10 | - | - | - | G |
| 7 (36-40f) | 16-20 | 12.18° / 14.56° | 0.40 / 0.32 | spots<sup>2</sup> | VisioCoach | - | G |
| 8 (56-60f) | 16-20 | 20.00° / 19.48° | 0.50 / 0.40 | - | - | - | G |

During the trials, participants were wearing Pupil Labs Invisible eye tracking glasses [27]. The device provides a 0.5° accuracy and a 200Hz refresh rate [27] according to the technical specs provided by the seller. Tonsen et al. [28] find that the mean bias of gaze-estimation ranges from below 0.5° up to 2.5° based on the VF region, with mean sample errors of 5° to 6.5°. Inertial measurements for tracking of head rotation use Madgwick’s algorithm [29], but no specifics are given about their accuracy and precision. Timestamps for start and stop of each trial were measured and automatically stored using a custom smartphone application built with the Unity3D game engine.

#### Experiment execution

Each session was initiated with four unmeasured trials to familiarize the participant with the task, the types of existing obstacles and the shape of the walking corridor. After that, 20 measured trials were done. Details on the measured parameters within these trials are found in Measurement parameters.

Prior to each trial, the experimenters positioned the obstacles according to one of 20 different layouts. Once the obstacle course was set up, the participant was guided to the starting position and directed to face forward towards the opposite side of the obstacle course. At this point, the Pupil Labs eye-tracking device was activated using the corresponding smartphone app. During this process, the participant had the opportunity to visually explore the scene, although only the initial third of the obstacle course was visible from the starting point due to the privacy screens. When everything was prepared and the participant confirmed their readiness, a second smartphone was used to start a timer and simultaneously play a sound cue, signaling the participant to start. While the participant navigated through the obstacle course, an experimenter followed them at a distance of approximately 3 meters, monitoring for any collisions that occurred during the trial. Upon reaching the designated goal area, the timer was stopped, accompanied by a second sound cue to indicate the completion of the trial.

During the session prior to the start of the gaze training, which could be either the first or second session, depending on the order of training and control phase, participants were introduced to the VR device and gaze training application. They were given the opportunity to familiarize themselves with the controls of the VR device and were guided through a tutorial of about 30 minutes duration in which the three visual tasks as well as the interface navigation within the gaze training application are explained. During this explanation, participants were also introduced to the suggested Scanpath pattern.

#### Measurement parameters

Two sets of measurement parameters were acquired as part of this study. The first set consists of the results of the real-world obstacle course trials, which were acquired during the in-person sessions. The second set consists of the performance and eye tracking information measured within the gaze training application during the four-week training phase. The following list summarizes all measurement parameters that were considered in the evaluation of this work:

#### Real-world obstacle course measurements

- **Trial Duration** Trial duration in the real-world obstacle course trials describes the time required by the participant to move from the starting position to the designated goal area. It was measured with the custom smartphone application described in Experimental setup.
- **Collisions** This parameter describes the number of obstacle collisions that occurred during a trial. Collisions were visually observed and documented by an experimenter who was closely following the participant during the trials. Collisions were categorized into two types: ‘Full collisions’ referred to frontal impacts where the obstacle was visibly and audibly struck - typically by the participant’s foot -, requiring a complete adjustment or re-evaluation of the movement path. ‘Light collisions’ referred to situations where obstacles were grazed or lightly touched, without impeding the participant’s motion.
- **Gaze Direction, Dynamic Field of View (DFoV) and Scanpath similarity** Using Pupil Labs Invisible eye tracking glasses, both the direction of gaze relative to the head and the orientation of the head itself are measured during the real-world obstacle course trials. Based on these two parameters, the Dynamic Field of View (DFoV) is calculated. The DFoV considers the expansion of the observed visual area through gaze movements. Essentially, it measures the average visual area that a person is able to observe over a fixed duration, considering both their head and eye movements as well as the size of their overall VF. For example, a DFoV over three seconds describes all the visual area that was covered by the participant’s limited VF at any point of time during the last three seconds. Then, by applying a rolling window of three seconds length over the entire duration of the trial, the average DFoV of this trial can be determined. The DFoV is reported and evaluated as a percentage change, showing how much the DFoV increased or decreased over the course of the training or control phase. The gaze direction is also used to determine saccades during trials which are required for the calculation of the Scanpath similarity value. Saccades are defined by a gaze movement speed of *>* 50°*/s* (based on Gibaldi et al. [30]) with a minimum saccade duration of 20*ms*.

#### Gaze training measurements

- **Target Tracking Task Performance** The performance of the Target Tracking Task - the task in which participants had to track and select a number of marked targets - was evaluated based on the number of incorrectly selected targets per trial. Each trial’s performance was measured on a point scale, where trials with no incorrect targets selected were rated with two points, and trials with one incorrect target equated to one point. However, due to the gradual change in difficulty levels of the task - described in Training tasks - rating each trial in the same way would not result in a good approximation of a participant’s total performance, as higher difficulty levels are likely to result in lower success rates. For the Target Tracking Task, the main factor that influences the difficulty of a task is the number of marked targets that must be tracked simultaneously. It is not feasible to compare a trial with only two marked targets to a trial with three or even four marked targets. Thus, to achieve a balanced approximation of task performance, only trials with four marked targets were considered, which is 45.1% of all measured trials.
- **Search Task Performance** In the Search Task, the base performance can simply be measured as the number of targets found and selected during the fixed 20-second time period of a trial. However, this again does not consider the change in difficulty level, which results in a larger or smaller area that has to be scanned to find the targets. To account for this, the Search Task performance score was adjusted based on the size of the search area in which targets would be placed, such that *P_adj_* = *n ∗* (*w_area_ ∗ h_area_*), where *P_adj_* is the adjusted performance score, *n* is the number of targets found and *w_area_* and *h_area_* being width and height (in visual angles) of the search area. In other words, to achieve the same performance score in a search area four times larger, the participant would have to find four times fewer targets.
- **Navigation Task Performance** The performance of the Navigation Task consists of two measurement parameters, both of which are reported on separately. The first parameter is the trial duration, which is the time taken from start to finish of the navigation course. The second parameter is the number of collisions during a trial. The layout of the obstacle course did not change with varying difficulty levels, making trials of different levels of difficulty more comparable to each other than in the other two tasks.
- **Gaze Direction and Dynamic Field of View** Similar to the real-world course trials, both head-centered gaze direction and head rotation were measured in all three visual tasks of the gaze training, using the VR headset’s built-in Tobii eye tracking device. This data was used to calculate the DFoV.

It must be noted that the performance scores calculated for the visual tasks of the gaze training are just an approximation of a participant’s actual skill level at different stages of the training, and are influenced by the methods that are applied to consider and eliminate the impact of varying difficulty levels on the performance.

#### Questionnaire

Five times during the training phase - following the initial training session and subsequently after every five training sessions -, participants were requested to complete a questionnaire to assess subjective ratings of enjoyment, motivation, stress, eye strain and other related factors. The questionnaire always featured the same questions, with seven of the questions following a 10-point Likert scale format and four questions allowing for free-form answers:

#### Questions to rank from 1 to 10

- **Enjoyment** - To what extent do you find each of the visual tasks enjoyable?
- **Motivation** - How motivated are you to improve your performance in each of the visual tasks?
- **Easiness** - How would you rate the ease of carrying out each of the visual tasks?
- **Stress** - To what degree do you experience stress while executing each of the visual tasks?
- **Eye Strain** - How straining is each visual task on your eyes?
- **Intuitiveness** - How intuitive is the use of the gaze training software?
- **Discomfort** - How much physical discomfort do you experience while wearing the VR headset?

#### Questions with free-form answers

- **Feedback for gaze training** - Which aspects of the gaze training application did you perceive as especially positive or negative?
- **Feedback for VR device** - Which aspects of the Virtual Reality headset did you perceive as especially positive or negative?
- **Feedback and suggestions** - What changes or improvements would you like to see implemented in the gaze training application?
- **Technical issues** - Did you encounter any technical issues during the training? If so, please describe.

### Evaluation process and statistical methods

In the real-world obstacle course tasks, four effects were tested for both training- and control phase.

- **Trial duration** *∼* **Pre-Post Condition** Using a Linear Mixed Model (LMM) with the trial duration as dependent variable and the pre-post-condition - meaning whether a trial was carried out in the session before or after the respective phase - as fixed factor, we can test whether there are significant changes to the trial duration after training phase or control phase, respectively. In addition to the fixed effect, participants were included as a random factor in the model, considering both random intercept (to consider different innate skills) and random slope (to consider different learning rates). Since trial duration results were not normally distributed and instead followed a right-skewed distribution, a logarithmic transformation was applied to the data to better meet the requirements of a LMM. A QQ-plot for the results with logarithm taken is found in Appendix C (S1 Appendix).
- **Collisions** *∼* **Pre-Post Condition** The collision parameter does not meet the requirements of a standard LMM, as its values are discrete count data, rather than continuous and normally-distributed. In addition, data was highly zero-inflated, with more than half of all trials (64.0%) showing zero collisions. Thus, to test the effect of each phase on the number of collisions in the real-world task, we applied a negative binomial Generalized Linear Mixed Model (nbGLMM) which is suited for this type of data [31, 32]. As before, the pre-post condition was considered as a fixed effect and participants were considered as a random effect, with one model testing the effects over the training phase and a second model testing the effects over the control phase.
- **Dynamic Field of View** *∼* **Pre-Post Condition** The DFoV was found to follow normal distribution quite well (the QQ-plot is found in Appendix C (S1 Appendix)) and data is continuous, allowing the use of a LMM with no required transformation. As described for the previous effects, the pre-post condition of each phase was once again used as fixed factor in the LMMs, participants were considered as a random factor.
- **Scanpath similarity** *∼* **Pre-Post Condition** The Scanpath similarity was evluated fully analogous to the DFoV. The QQ-plot, indicating the normal distribution of the data, is once again found in Appendix C (S1 Appendix). While the order of the obstacle trials was changed between sessions, each of the 20 obstacle trial layouts was used exactly once per session. This ensures that the effect of different layouts on the performance within the obstacle course does not have to be considered in the statistical models. Regarding the results of the Virtual Reality gaze training, it was modeled and analyzed how the task performance as well as the DFoV in all three visual tasks changes over the course of the training.
- **Target Tracking Performance** *∼* **Training Session** As described in Measurement parameters, the performance in the Target Tracking Task is based on the number of incorrectly selected targets at the end of a trial. To measure this, a point scoring system was employed, where a score of 2 points was assigned for trials with zero errors, 1 point for trials with one error, and 0 points for trials with two or more errors. This means that the Tracking Task Performance can be treated as count data, and thus a Generalized Linear Mixed Model (GLMM) was employed for the analysis. Fixed factors of the model are the training session number (from 1 to 20) as well as the number of the current trial within the training session, as both can be assumed to have an influence on the task performance. Once again, participants were considered as random factor with both random intercept and random slope.
- **Search Task Performance** *∼* **Training Session** Search Task Performance was measured as the number of stationary targets found in a 20 second interval. QQ-plots found it to roughly follow normal distribution, making the use of a LMM suitable for analysis. As before, fixed factors of the model included training session and trial number, participants are considered as random factor.
- **Navigation Trial Duration** *∼* **Training Session** Similar to the real-world obstacle course trials, the trial duration of the Navigation Task trials was found to be right-skewed, thus the logarithm was taken for the analysis. A LMM was employed analogues to the previous analysis of the Search Task Performance.
- **Navigation Trial Collisions** *∼* **Training Session** The number of collisions per trial can be treated as zero-inflated count data, similar to the collisions in the real-world obstacle course. This indicates the need for a nbGLMM, where training session and trial number are treated as fixed factors, participants as random factor.
- **Dynamic Field of View** *∼* **Training Session** The DFoV was analyzed the same for all three visual tasks using a LMM. No transformation was required, as DFoV followed normal distribution in all three tasks according to QQ-plots. Following the previous models, the DFoV was tested against the training session and trial number as fixed factors, with participants being considered as random factor.

The alpha level that determines the threshold for statistical significance was chosen as 0.05 for all models. All errors are reported as the standard deviation of results. Analysis was done using R and the RStudio graphical interface with the nlme and lme4 library. The detailed models and results of the statistical analyses can be found in Appendix C (S1 Appendix). The results of the questionnaires are reported on qualitatively, as the number of samples is too low for statistical analysis.

For the real-world obstacle course results, it must be mentioned that 61 out of 480 measured trials did not include complete gaze-tracking data (25 of these trials included head-centric gaze data but no head rotations, 36 trials were missing both eye- and head-tracking data) and thus had to be discarded from the analysis of DFoV. This loss of data was likely caused by a shaky contact of the eye tracking device and was only detected late in the experiment phase. The data loss affected three sessions in particular: The eye-tracking data was lost or incomplete in 15 out of 20 trials in the second session of participant 4, 18 out of 20 trials in the second session of participant 6, and all 20 trials in the first session of participant 1. Thus, for participant 1, no results for the change of DFoV over the control phase could be evaluated.

### Deviations from original study protocol

This chapter lists the deviations of the actual methods from the original study protocol found in the supporting files (S2 File).

- The initial proposal outlined the use of a Fove-0 VR headset for the study. In the time between the ethics application and the start of the study, new VR devices became available that were better suited for at-home training, notably the Pico Neo 2 Eye, which ultimately became the chosen hardware for this study. The main advantage of the Pico Neo 2 Eye compared to the Fove-0 is its self-contained hardware, as it does not require any external hardware setups.
- The protocol allocated a total of 30 RP patients for the study, divided into two groups of 15 patients each: A training group and a control group. During the patient acquisition phase it became clear that the number of RP patients interested in study participation would not allow for this study population size. Thus, the design was changed to a crossover study, with all patients carrying out both training and control phase in randomized order, as is described in Experimental study. Additionally, the protocol provided for inclusion of an additional group of visually healthy patients as control. This plan was discarded because the low relevance of the results to be obtained from this group would not have justified the additional time and material effort.
- The setup of the real-world obstacle course used in the study deviates from the one described in the protocol. The protocol outlined a 68m long and 1.3m wide corridor. At the time of the study, no location was available that would have allowed for a course of these dimensions. Thus, the course was adjusted to the dimensions described in Experimental setup.
- The protocol planned for a mandatory ’eye motion’ task as part of the gaze training. As was mentioned in Scanpath behavior and will be further addressed in Discussion, the part was instead included as a voluntary task, following the findings of a preliminary study focused on the effects of Scanpaths in gaze training [25].
- In addition to the real-world obstacle course, the protocol provided for a performance evaluation in a realistic city environment within the virtual world. This plan was discarded because the development of a realistic 3D city environment would have been beyond the available time and expertise for the study.
- Lastly, the methods for statistical analysis were changed. The original protocol outlined the use of t-tests and mixed model repeated measures ANOVA analysis. Over the course of the study assessment, it was decided that Linear Mixed Models as well as negative binomial Linear Mixed Models are more suitable for the statistical evaluation of the acquired data.

## Results

### Real-world obstacle course

A total of 480 real-world obstacle course trials were absolved, split among 8 participants with three sessions each. The raw result tables for the trials are found in the supplementary material of this work. Fig 8 shows the results for navigation performances and the DFoV of the eight participants.

**Fig 8.**
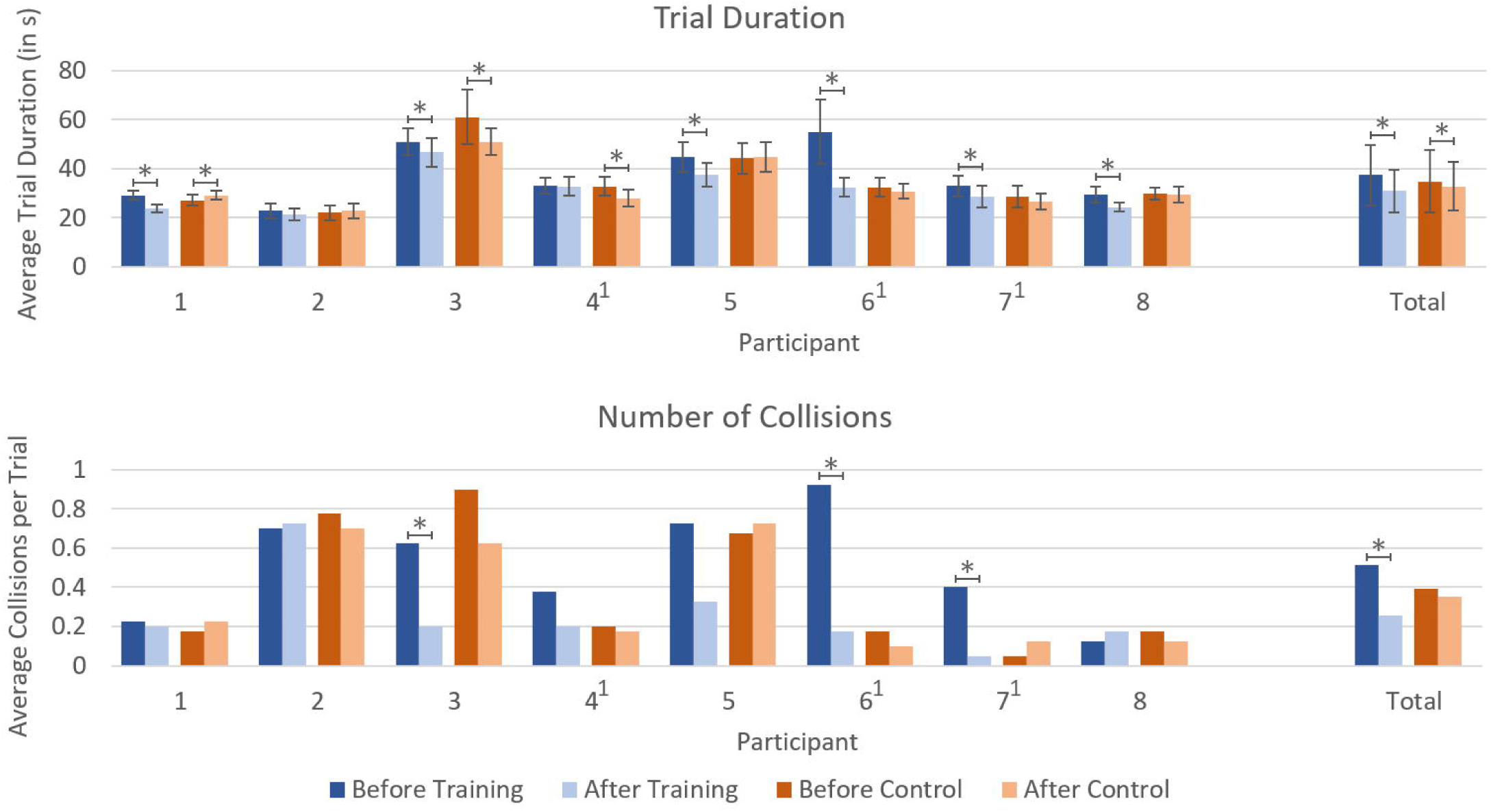
Obstacle course results. Participants’ performance in the real-world obstacle course trials before and after each of the two phases. * indicates significance (*p <* 0.05). ^1^ marks participants who carried out the training phase before the control phase; all other participants started with the control phase and carried out the training phase afterward.

#### Navigation performance: trial duration and number of collisions

After the training phase, participants displayed a significant improvement in trial duration by 17.0% (*p <* 0.001) compared to the performance before the training, decreasing the average trial duration from 37.2 (±12.3) seconds to 30.9 (±8.68) seconds. In comparison, after the control phase the average trial duration was found to have improved by 5.9% (*p* = 0.003), from 34.8 (±12.7) seconds to 32.7 (±9.87) seconds. The average number of collisions per trial decreased by 50.0% (*p <* 0.001) after training, from 0.513 collisions per trial to 0.256 collisions per trial. Meanwhile, after the control phase the average number of collisions per trial improved only by 10.4% (*p* = 0.609), from 0.391 to 0.350 collisions per trial. Overall, the results suggest that the training phase was significantly more effective in improving the average trial duration and reducing the number of collisions compared to the control phase.

Out of the four obstacle types in the real-world obstacle course - horizontal box, vertical box, stepping obstacle and hanging obstacle - the type that caused most collisions is the stepping obstacle with a total of 76 full collisions and 53 light collisions in all 480 trials. However, it is shortly followed by the hanging obstacle at 75 full collisions and 52 light collisions. Considering that each obstacle course layout features six stepping obstacles, but only three hanging obstacles, it can be stated that the hanging obstacles pose the highest risk for collisions. Very few collisions were tracked for the horizontal and vertical boxes, with 6 full collisions and 8 light collisions for horizontal boxes and 3 full collisions and 13 light collisions for vertical boxes.

#### Visual Performance: Dynamic Field of View and Scanpaths

Using the eye-tracking data collected during the real-world obstacle course trials, it is possible to evaluate how the average DFoV of participants - the visual area observed over time - changed over the course of training and control phase, as shown in Fig 9. On average, participants displayed an increase in world-centric DFoV of 4.41% (*p <* 0.001) after the training phase and an increase of 2.06% (*p* = 0.0835) after the control phase. However, only three of eight participants (1, 3, 6) show a notable increase after the training phase, with two participants (4, 8) showing decreases. When considering only head-centric eye movements, no significant change in DFoV is found for the average DFoV after either phase, with a change of -0.052% (*p* = 0.175) after training phase and 0.108% (*p* = 0.383) after control phase. While this suggest that the increase in DFoV originates mainly from a change in head movements, rather than eye movements, two of the three participants (3, 6) with notable increase in world-centric DFoV also show similar increase in gaze-centric DFoV.

**Fig 9.**
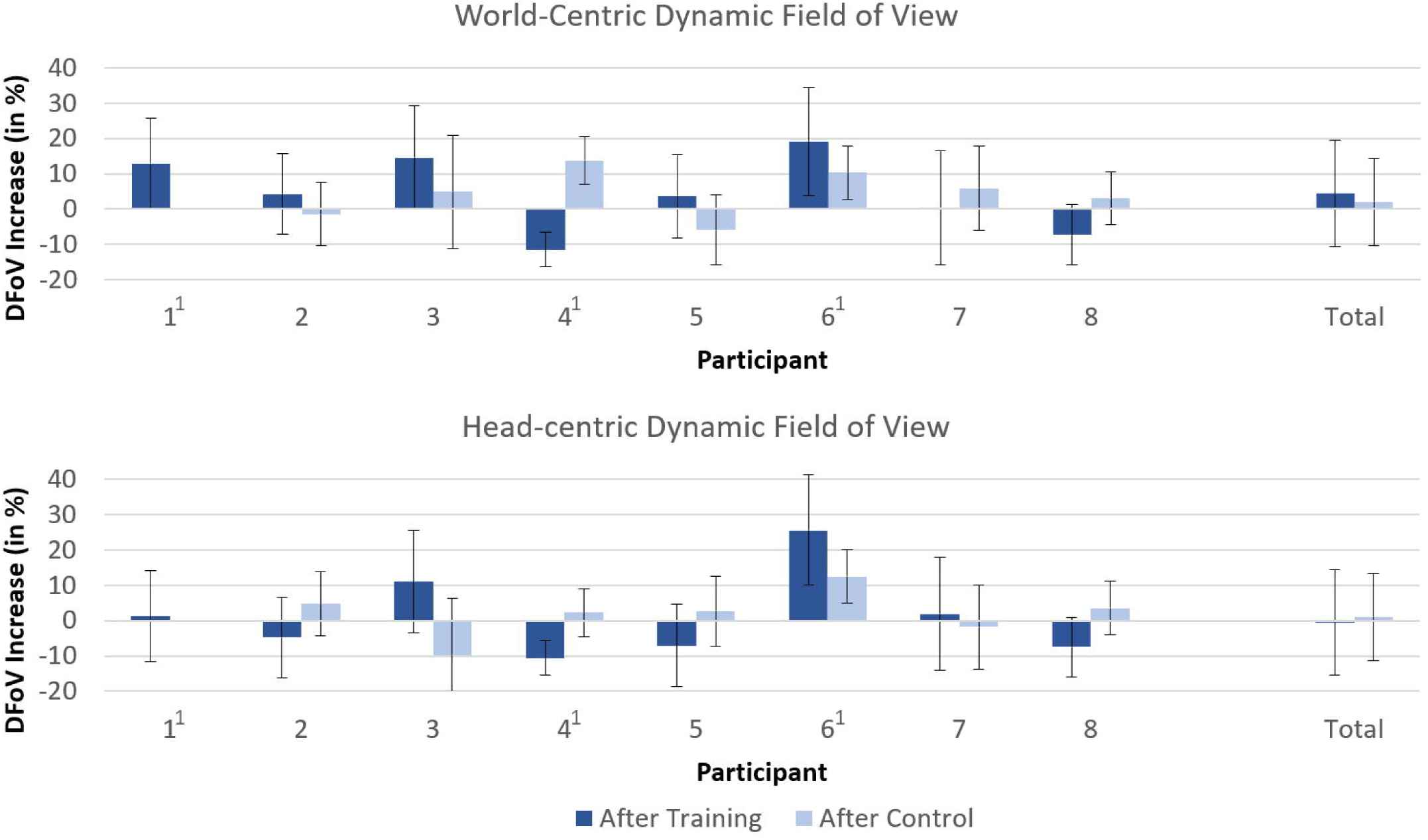
Dynamic field of view. Increase of the Dynamic Field of View in the real-world task after training phase or control phase respectively. Top graph shows the results based on world-centric gaze data (considers both head- and eye movements) whereas the bottom graph shows results based on head-centric gaze data (only eye movements, no head rotation considered). DFoV was calculated over a 3 second rolling window. ^1^denotes participants with incomplete or missing eye-tracking data.

The results of the evaluation of the Scanpath similarity - the value describing how closely the participants’ displayed gaze movements match the suggested Scanpath that was presented in Fig 4 - is shown in Fig 10. There is no significant increase found after either of the two phases (*p* = 0.168 for training phase, *p* = 0.147 for control phase), and the similarity values give no indication that participants were actively following the suggested Scanpath.

**Fig 10.**
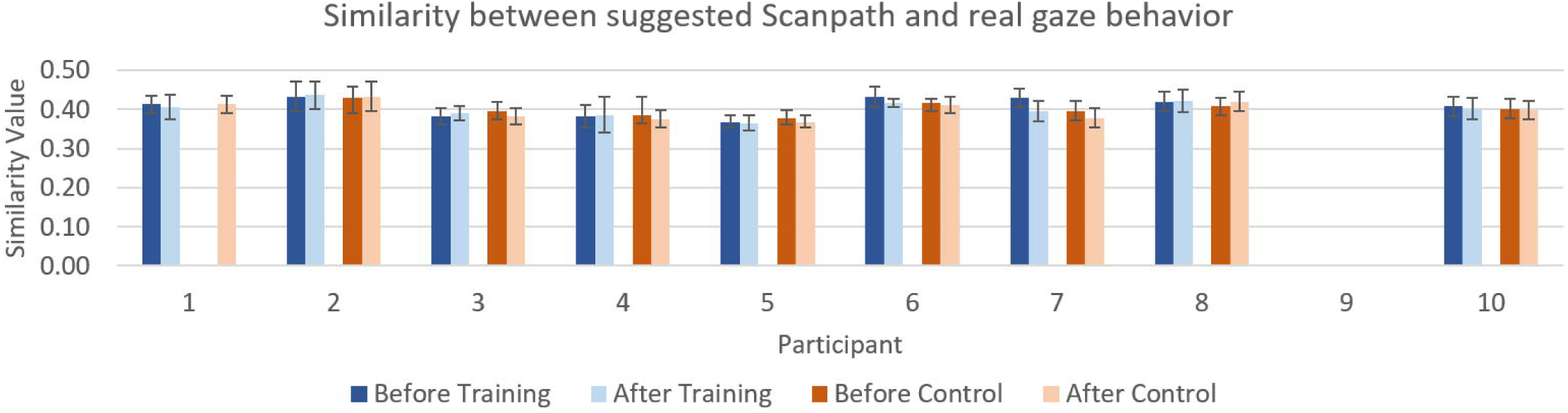
Scanpath results. The figure shows the average similarity between the gaze pattern displayed by participants and the suggested systematic Scanpath described in Scanpath behavior. Values between 0.3 and 0.5 are common for naive gaze behavior, whereas values between 0.6 and 0.8 can be expected when a subject is actively following the Scanpath.

### Virtual-reality gaze training

For the gaze training, a total of 3125 Target Tracking trials, 3205 Search Task trials, and 2583 Navigation trials were evaluated, distributed across the eight participants and *∼*20 training sessions per participant. Fig 11 shows the increase in visual task performance and DFoV within the virtual environment of the gaze training, shown as the percentage improvement from the start of training to the end.

**Fig 11.**
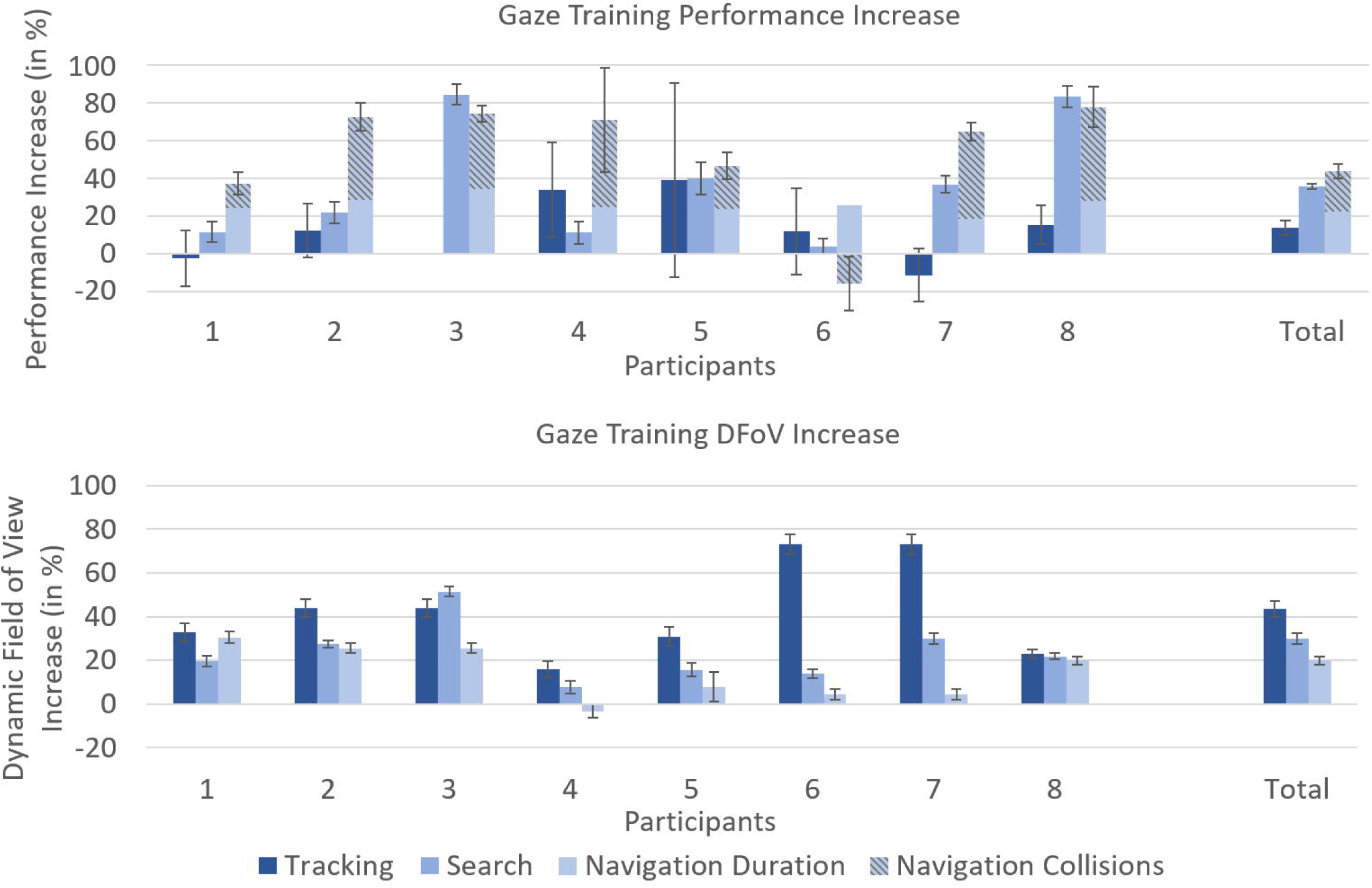
Gaze training results. Percentage increase in approximate task performance (top) and average Dynamic Field of View (bottom) within the VR training setup. Reported DFoV is head-centric and measured over a 3 second rolling window.

Most participants improved both in performance and in average DFoV in all three tasks. In the Search Task, the average task performance increased by 35.5% (*p <* 0.001). In the Target Tracking Task, performance increased by 13.9% (*p <* 0.0032). The average task performance in the Navigation Task increased by 44.3% for trial duration (*p <* 0.001) and 42.8% for collision avoidance (*p <* 0.001). Despite showing the lowest task performance increase, the Target Tracking Task evoked the highest increase in average DFoV over the course of training, with an increase of 43.4% from beginning to end of training (*p <* 0.001). The Search Task followed with an increase in DFoV of 29.9% (*p <* 0.001), and the Navigation Task resulted in the lowest average DFoV increase out of the three tasks at 19.8% (*p <* 0.001).

#### Questionnaire

The questionnaires filled by participants over the course of the gaze training give insight into qualitative results. Fig 12 shows the average results of all five questionnaires for the seven ranking questions related to the VR gaze training shown in Questionnaire.

**Fig 12.**
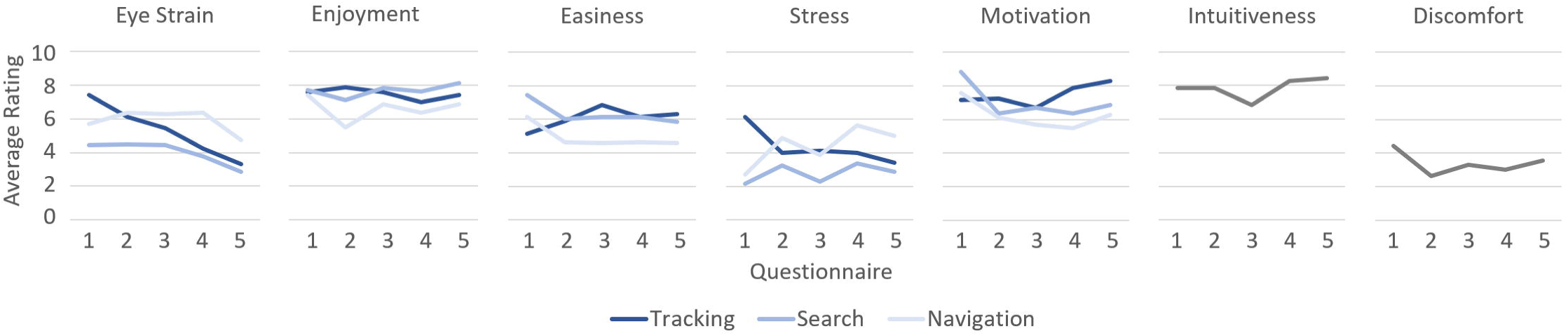
Questionnaire results. Average rating of different aspects of the gaze training. Questionnaires 1-5 were filled after 1, 5, 10, 15 and 20 training sessions. A rating of 10 equals ”very high”, a rating of 1 equals ”very low”.

Overall, the Search Task was rated most positively, with high enjoyment and motivation for improvement, as well as low perceived stress and low eye strain reported. On the opposite side, the Target Tracking Task was rated most negatively, with the lowest task enjoyment and motivation to improve upon previous results, and highest stress and eye strain reported out of the three tasks. Participants also gave feedback for the gaze training, both in the questionnaires and in the in-person training sessions. Some participants stated minor technical issues both with the VR headset and the developed software. Additional suggestions included more variety in tasks or task visualization, such as adding different backgrounds; As well as quality-of-life changes, such as more fluent turn animations for the moving targets in the Target Tracking Task, or a better indication before the application switches to the selection menu at the end of a trial.

The raw data results for the obstacle course trials, the VR gaze training, as well as the questionnaires are found in the supporting files (S1 File).

## Discussion

In this study, the effects of 10 hours of training with a VR based gaze training on the navigation performance in a real-world obstacle course task was evaluated in eight Retinitis pigmentosa patients. It is found that the navigation performance increase over the training phase significantly surpasses the natural learning effect found after the control phase, suggesting that Virtual-Reality based gaze training can improve navigation performance of patients with Retinitis pigmentosa. A small, but significant increase in DFoV is found after the training phase, which shows that the gaze training affects the participants’ gaze behavior even in a real-world task, once again suggesting the general feasibility of VR gaze training methods.

Analysis of the real-world obstacle course results for each individual participant reveals a significant variability in the impact of gaze training on the DFoV. Among the eight participants whose results were evaluated, three demonstrated a prominent increase in DFoV during the real-world task after training (Fig 9). Notably, two of these participants (participant 3 and 6) also exhibited significant improvements in both trial duration and collision avoidance (Fig 8), with the third participant (participant 1) showing a significant improvement only in trial duration, with no corresponding increase in collision avoidance. These findings imply a positive correlation between an expanded DFoV and improved navigation performance. However, it is worth noting that improvements in trial duration and collision avoidance were also observed in participants who did not experience any changes in DFoV following training. Participants 5 and 7 exhibited significant improvements in trial duration and noticeable improvements in collision avoidance, despite no discernible change in DFoV. Likewise, participants 4 and 8 - the only participants to display a decrease in DFoV after training - did not exhibit any negative effect on trial duration or collision avoidance, which would be expected when assuming a direct correlation between DFoV and navigation performance.

In summary, these findings suggest that an increased DFoV is likely to positively impact navigation performance, with no evidence indicating that an expanded DFoV could have a detrimental effect. However, the fact that trial duration and collision avoidance improved in some participants without changes in DFoV suggests the presence of other factors through which gaze training influences navigation performance. Participant 4, 5 and 7, for instance, all demonstrated improvements in the Navigation Task over the course of the training despite minimal changes in their DFoV (Fig 11). This aligns with the results obtained from the real-world obstacle course (Fig 8 and Fig 9), where participants also displayed improved navigation performance with no increase or even with decrease in DFoV. Based on these findings, it is plausible to speculate that the Navigation Task facilitated the development of spatial awareness in these and maybe other participants. It is also possible that through the Navigation Task, participants adapted to previously less familiar types of obstacles, such as obstacles hanging from the ceiling.

In Scanpath behavior, a previous study was mentioned [25]. Here, we assessed whether systematic eye movement patterns (or ”Scanpaths”) could assist people with tunnel vision condition in navigation and obstacle avoidance. For that, a gaze-contingent simulation of a 20° diameter tunnel vision condition was employed on visually healthy participants within a Virtual Reality environment, and the participants navigated through a virtual obstacle course while following different Scanpath eye movements in a supervised experimental setup. One of the tested Scanpaths - a mostly horizontal, serpentine scanning motion - was found to significantly reduce obstacle collisions by 32.9% and increase the DFoV by 8.9% compared to trials without systematic gaze movements. This came at the cost of a significantly lower movement speed, resulting in an average of 24.6% longer trial duration when following the Scanpath. The study concluded that the Scanpath has the potential to enhance visual performance in the presence of tunnel vision and suggests that introducing the Scanpath in gaze training for individuals with tunnel vision could have beneficial effects. However, it is important to note that the Scanpath study did not involve any real RP patients and no effect of the execution of Scanpaths in a real-world setting were evaluated in this study. Given the non-conclusive nature of the results of this previous study, the execution of the suggested Scanpath was included as a voluntary task for the gaze training presented in this work.

As the evaluation of the Scanpath similarity before and after the training phase indicates, the participants have not adapted this gaze pattern. This is also reflected in participants’ statements after the training. Six out of the eight participants (1, 2, 3, 4, 5, 8) reported that they did not follow any specific gaze movement strategies. The predominant reason stated was that they ”didn’t think of it” during the trials. Participants 6 and 7 reported to follow individual Scanpaths. Participant 6 described a radial Scanpath, moving the gaze in a small circle at first and then in a second, larger circle. She reported to have adapted this Scanpath strategy during the Target Tracking Task of the gaze training and is now using it successfully even in everyday life. Participant 7 reported to follow a cross-like Scanpath, moving the gaze vertically from bottom to top, then left to right. However, she reported to have adapted this gaze pattern only for the real-world obstacle course trials and not during the gaze training. Overall, the outcome implies that systematic Scanpaths are not easily and voluntarily adapted in practical application by the majority of patients. Still, the fact that participant 6 - as one of two participants reporting to follow a Scanpath during trials and the only participant stating to also follow the Scanpath in everyday life now - displayed the highest increase in both DFoV and navigation performance in the real-world obstacle course suggests further research towards the training of individualized Scanpaths in people with limited VF.

The feedback received by participants was overall positive. During the study, six participants (participant 3, 4, 5, 6, 7 and 8) expressed their interest and willingness to continue the gaze training if it becomes available, though participant 8 specified she would prefer to only do the Search Task if training continued. Two participants mentioned to have recommended the training software to friends and acquaintances, and two participants stated to regularly notice improvements in visual and navigation performance in everyday life since the training phase started. Questionnaire results support the overall positive reception of the gaze training, with consistently high ratings in task enjoyment (average 7.26/10) and intuitiveness and ease of use of the software (average 7.85/10). Participants reported some eye strain in the beginning of the training (average 5.86/10) which decreased towards the end of training (average 3.61/10).

To our knowledge, only two other studies were published that investigate the effect of gaze training on the real-world navigation performance in Retinitis pigmentosa patients [13, 33]. It must be noted that quantitative comparison between the results of different gaze trainings is feasible only to a very limited degree due to the different experimental setups in which they were acquired. In their study investigating the effect of a computer display based gaze training application on navigation performance, Ivanov et al. developed a gaze training that consists of an exploratory search task very similar to the Search Task of our work. It was found that RP patients displayed a significant increase in their preferred percentage walking speed by *∼*6% in a real-world obstacle course after a six-week training period (total of 15 hours), with no significant improvements in obstacle avoidance in the trials. It was already suggested by Ivanov et al. that the application of VR devices may impact the effect of gaze training positively. Considering the notably larger improvements found after the VR training - compared to those found after training with a screen-based setup - it can be assumed that VR based gaze training shows higher potential to improve navigation performance in real-world tasks.

However, the VR based setup also shows some limitations: As mentioned in Study population, two participants, in addition to the eight who completed the study, withdrew from the study at an early stage. The primary reason for their discontinuation was directly associated with the use of a VR headset for the training sessions. The first of the two patients reported an increase in migraine attacks when carrying out the training. Virtual Reality is known to cause motion sickness or headaches in some users [34, 35] and it is thus likely that the use of the VR headset did have an effect on the increase in migraine attacks. It was decided to stop the study participation after three training sessions to avoid any risk and discomfort for the participant. The second participant who discontinued the study displayed severe difficulties in navigating within the VR environment when first being introduced to the gaze training. They reported feeling completely disoriented and unable to complete the tasks on their own, and it was thus decided that carrying out the training in an unsupervised at-home scenario would not be feasible.

A common feedback from participants was that the starting difficulty of the Navigation Task as well as the threshold to advance to higher difficulty levels was too high. Three out of eight participants were not able to advance to a higher difficulty level over the entire duration of the study. They still displayed improvements in performance, but they did not reach the threshold - calculated based on a combination of trial duration and number of collisions - at which the difficulty level would increase. This suggests lowering the starting difficulty of the Navigation Task by decreasing the length of the course as well as the number of obstacles.

Contrary to the Navigation Task, the Target Tracking Task has a low entry difficulty, however participants reported a steep increase in both difficulty and resulting stress at higher difficulty levels. The main factor is the increase of targets that must be tracked simultaneously in order to be successful in the task. Tracking two targets did not prove a big challenge for any of the participants, however once a third target is introduced, difficulty and stress are drastically increased, and none of the participants were able to consistently track more than three targets at the same time. This lead to a stagnation at the difficulty levels around the threshold between three and four marked targets for many participants. To avoid this issue, it may help to limit the number of marked targets to three and instead purely focus on the increase of other difficulty parameters, such as the movement speed of the targets or the area in which targets are free to move around.

Despite these limitations, the developed gaze training shows very promising results, and the use of Virtual Reality as a medium for gaze training seems feasible. Based on feedback from the participants of the study as well as some general impressions of the training process, several changes will be made to the software.

- Difficulty levels will be adjusted to allow for an easier start as well as a more gradual increase of difficulty. In addition, users will have the option to manually change the current difficulty level.
- To allow for fully unsupervised usability of the training software, a tutorial will be created to introduce users to the three visual tasks.
- Currently, the gaze training software only works with the Pico Neo 2 Eye VR headsets that were used for this study. In order to increase accessibility of the training task, compatibility with other popular VR headsets, such as the HTC Vive series or the Oculus Rift and Oculus Quest will be targeted, allowing everyone with access to one of these devices to test the gaze training.
- Lastly, some minor quality-of-life changes and stability improvements will be implemented to address some of the feedback points from the participants.

The gaze training is currently published as a ’work-in-progress’ open-source software on GitHub [36]. This will allow everyone with access to one of the supported VR devices to test and use the gaze training for free once the changes are implemented.

## Conclusion

The results after four weeks of training with the developed gaze training software are promising, showing that Virtual Reality gaze training has the potential to improve the visual and navigation performance of people living with Retinitis pigmentosa in real-world tasks. The majority of participants reported the training software - along with the Virtual Reality device - to be intuitive and easy to use, making it suitable for at-home training with no supervision and with minimal introduction time. However, while VR proves to be a viable medium for a gaze training tool, it can also act as an entry barrier for people being susceptible to motion sickness or people facing difficulties with orienting and navigating in virtual environments. Still, the developed gaze training shows potential to have a significant positive impact on real-world navigation performance and is currently available as work-in-progress open-source software (link: https://github.com/ANCoral05/VR-GT-Virtual-Reality-Gaze-Training).

## Supporting information

S1 Consort

## Data Availability

All relevant data are within the manuscript and its Supporting Information files.

## Supporting information

**S1 File. Measurement data** The raw measurements for the real-world obstacle course, the three gaze training tasks as well as the participant questionnaires.

**S2 File Study protocol** The original study protocol as approved by the ethics committee.

**S1 Video. Visual training tasks.** A screen recording of the three visual training tasks of the gaze training.

**S2 Video. Obstacle course trial.** Example of one of the trials in the real-world obstacle course, captured by the scene camera of the Pupil Labs Invisible eye tracker.

**S1 Appendix. Appendix** Additional information regarding the Scanpath evaluation, VF data and the results of the statistical methods applied in this study.

**S1 CONSORT checklist**

## Acknowledgments

We would like to express our sincere appreciation to the Deutsche Forschungsgemeinschaft (DFG) for funding this study. For her indispensable supervision, guidance, support, and expertise, many thanks go to Enkelejda Kasneci of the Technical University of Munich (TUM). We also thank Gabriele Roever and Stefan Küster of Pro Retina Baden-Württemberg for their advice and aid in acquiring participants for this study. Further gratitude goes to Nadine Wagner of the Tübingen city administration for her assistance in securing a location for the experiment. Lastly, we extend our thanks to all our study participants for their contributions and feedback.

## Notes

### Competing Interest Statement

The authors have declared no competing interest.

### Clinical Trial

Registry: German Clinical Trials Register (DRKS) ID: DRKS00032628 URL: https://drks.de/search/de/trial/DRKS00032628 The registration was done retrospectively, as the study was originally considered as non-interventional observation study, not as clinical trial. Prompted by later feedback, this decision was reconsidered and the study was registered.

### Funding Statement

This work was supported by the Deutsche Forschungsgemeinschaft (DFG), URL: https://www.dfg.de/en/ (Grant: DFG IV 167/5-1 to II). The funder did not play any role in study design, data collection and analysis, decision to publish, or preparation of the manuscript.

### Author Declarations

Ethics committee of the Institutional Review Board of the Medical Faculty of the University of Tuebingen gave ethical approval for this work in accordance with the 2013 Helsinki Declaration. The project number is 628/2018BO2. Patients were recruited in the time from June 9, 2022 to November 22, 2022. All participants signed informed consent forms.

