## Supplementary material for "Influence of open-source virtual-reality based gaze training on navigation performance in Retinitis pigmentosa patients in a crossover randomized controlled trial": S2 File

### B PROJECT DESCRIPTION

**“Improving visual performance in retinitis pigmentosa by new non-invasive personalized and adaptive training.” by Dr. Iliya V. Ivanov, University of Tuebingen, Tuebingen**

#### 1 State of the art and preliminary work

*State of the art*

The aim of this proposal is to study the pathological visual behaviour in patients with retinitis pigmentosa (RP) and to implement new methods able to improve residual visual performance. The project goal is directly related to the overarching topic of the first workshop for early career investigators "Neurosensory Systems Prevention-Regeneration-Restitution" with the focus on disorders of eye and ear funded by the German Research Foundation. In particular, the goal is aimed at *improving* immediate condition and relates to vision regeneration and restitution. To achieve this goal, and to alleviate the severe navigation problems that RP patients suffer in daily life, new personalized training protocols are designed and tested. The importance of these new specific eye-movement training protocols is crucial in order to optimize residual vision in retinitis pigmentosa (RP). Up to date, visual loss due to RP, which is the most common hereditary retinal diseases (retinal dystrophy) affecting 1 in 3000 individuals (1), is irreversible (1-3). RP is a retinal degenerative disease that typically causes a severe concentric visual field (VF) loss, also known as tunnel vision. It is characterized by progressive loss of rods, followed by loss of cone photoreceptors (1-3). Thus, at early RP stages, peripheral vision is damaged, while central vision is relatively intact with good visual acuity. Later RP stages, however, impact central vision and acuity. Typically, over a period of 15 to 20 years, RP patients’ visual field widths decrease to as little as 4 degrees and then, within several more years, total blindness occurs.

*The role of the peripheral vision in binding the global structure of the environment*

Peripheral vision, despite its low resolution, is important for creating and updating an accurate representation of spatial structure for navigation (4-6). Healthy observers process complex scenes easily and are able to integrate effortlessly the information about the world over time. This integration of information over time may be achieved in two ways. First, observers move their eyes around a scene to overcome the resolution problems caused by the peripheral vision. Secondly, observers shift attention around the scene to process various objects like possible obstacles. As a result, over time, what has been seen needs to be analysed with relation to what is being seen. That is, the peripheral visual field provides the spatial framework in which to place the current and following fixations (6). Fortenbaugh and colleagues (6) found that eye and head movement behaviour did not change when normal field of view (FoV) of healthy subjects was narrowed to 40, 20 or 10 deg. They also showed, however, that the loss of input to the peripheral visual field led to systematic distortions in representations of object locations in space: decreasing FoV size led to heterogeneous compressions of estimated distances in the scene. These findings were interpreted as an evidence that one of the functions of the peripheral vision is to deliver information about the global framework of the environment within each fixation. Further, there are implication from the boundary extension literature (7-9) that the visual system uses heuristics about the stability and continuity of an external environment to extrapolate beyond the limits of current sensory input, at a given fixation. Thus, taken all these studies together (4-9) suggests that while the online perception of a stable scene would not differ when seen through different FoV sizes, the resulting spatial representations may contain unique distortions, where smaller FoVs would lead to larger extrapolations of the space over time and thus failing to incorporate changes in the true global structure of the environment that could be used as landmarks when trying to safely navigate and avoid possible obstacles. Therefore, RP, which is a condition characterized by the lack of peripheral vision in its early stages impairs mobility (4).

It is known that due to the lack of peripheral input, the most frequent mobility problems in people with tunnel vision include walking more slowly than normally sighted people, fear of falling, collisions with objects and bumping into other people (1-4). Additionally, at the early stages of RP, patients also complain and experience difficulties while navigating in conditions including changes in elevation and instantaneous change of light levels (4). Also, these patients have severe difficulties performing in dim light, a condition commonly known as night blindness. This type of tunnel vision hinders daily activities and makes tasks that otherwise seem effortless, extremely challenging. Evidence about navigation problems in RP comes from our preliminary work, where we demonstrated that on average RP patients performed worse as compared with healthy subjects in a task to navigate in a hallway and avoid stationary obstacles: RP patients walked slower, and hit more obstacles (4). This finding relates with the main problem of the RP patients, that is, their inability to create proper spatial reference frame of the environment in which to place the content of the current fixation. Think of a giant puzzle, where patients are unable to see the global picture, or the boundary of the puzzle, within which all pieces of information acquired during consecutive fixations need to be positioned. Therefore, our main goal of this proposed research is to provide the patients with a neural-plasticity training that helps them to reconstruct the global picture with their limited VFs.

While gene therapies in RP may promise future sustained functional rescue (10), contemporary medical and therapeutic treatment are still limited and currently visual loss cannot be reversed. Gene therapies are among the most promising future treatments, since RP is caused by defects in certain genes (11-14). However, clinical trials have demonstrated only temporary improvement and in general, as a major issue that impacts the implementation and testing of any type of therapy is the timing of treatment (14). For instance, in preclinical studies, gene therapies are almost always delivered before the onset of the cell degeneration (14). On the other side, it has been suggested that in RP treatment at later stages might be hindered by a “point of no return”, beyond which damage is irreversible (14). Since most neural degenerative diseases are diagnosed after the onset of the cell loss, when the retinal damage is irreversible, an important research priority, therefore, is to provide and validate personalized training tools able to improve patients’ immediate condition.

Currently, visual performance can be recovered, at least partially, by the means of traditional optical devices (15). However, recovery in the lost visual functionality is usually at the cost of other visual faculties. For instance, optical systems are not flexible and while magnifying devices ultimately increase resolution, this is usually achieved at the cost of notable decrease in the visual filed. Thus, magnifiers further reduce the already small VF and hence they do not improve mobility. Additionally, a magnified letter in a small VF is not helpful in a reading task. Yet another approach to recover visual performance, is to use the innate neural plasticity and provide specific eye-movement training protocol that helps patients to naturally “expand” their restricted VF (4): It is known that brain neural plasticity may involve modifications in overall cognitive strategies to successfully cope with new challenges. This involves attention and behavioral compensation (16, 17) for the recruitment of new/different neural networks (18-21), or for changes in strength of connections in existing networks in charge for the execution of a particular movement, language, vision or hearing task (22, 23). These changes may be demonstrated in animals and humans and are measured *in vivo* and *in vitro* at the cellular level as changes in membrane excitability, synaptic plasticity, as well as structural changes in dendritic and axonal anatomy (24, 25). At functional level, behavioural compensation due to neural plasticity may be demonstrated as improved performance after training of new strategies to perform a challenging task. For example, our preliminary research (4) demonstrated that explorative saccade training (EST), a non manipulated, eye-movement training that rely on the natural course of strategy acquisition, selection and refinement, led to sustained improvement of visual performance related to daily life in RP patients. The rationale for this type of training is based on two established facts: It was shown in a variety of paradigms that scanning eye movements are led by attention (15, 26). Secondly, there are two mechanisms of attention that may be relevant here: A “sustained” component that is slow, voluntarily controlled and does not depend on visual input, and a “transient” component that is fast, reflex-like, and depends on visual input (27, 28). This is why transient attention is based on “bottom-up” processing of exogenous information, while sustained attention is a mechanism that uses goal-oriented “top-down”, endogenous commands (27-35). For example, salient stimuli can attract transient attention, even though the subject had no intention to attend to these stimuli (29, 30). On the other hand, if visual input is missing or compromised, as e.g. by a retinal disease like RP, the transient and stimulus-driven mechanism does not work anymore. In this case, goal-directed “sustained” attention (27-35) can be voluntarily directed to objects and features in the seeing visual field, and even to locations in unseen regions in space. Thus, we can ask subjects to make eye movements towards targets in their non-seeing visual field using sustained (top-down) attention, which is slow and consciously controlled, independent of visual input (4).

Our preliminary research findings, described into detail below, clearly demonstrate the potential of neural plasticity to improve and to recover the patients’ visual performance, which was assessed via mobility tests and evaluating eye movement behaviour during walking. We implemented the neural-plasticity training in RP as a simple computer based visual search task of static targets, presented among distractors on a screen. In particular, we implemented EST as a saccadic search task with the aim to improve visual search outside the seeing VF, improve the use of the total field of gaze and thus to improve the awareness of the size of the visual field defect. It is assumed that sustained attention in this case was guided serially to find one target at a time, at the different retinal loci. Importantly, during training, guidance related to the possible location of the targets was not provided to the patients, and neither the eye movements were recorded. That is, unlike in daily life, the training task was restricted to simple random search task, targets were static and patients did not move. During the course of the training, patients developed new eye movement strategies to find targets presented in their non-seeing visual filed. Visual performance of patients was tested before and after training on a task similar to daily life: to navigate and avoid obstacles in a controlled laboratory condition. It was shown that the patients usually performed better after training, were able to maintain improvement at follow-up (six weeks after training) but restored visual performance was far from normal (4). Since RP is progressive and visual function declines over time, we envisage that patients should keep practice, albeit at a slower rate, the neural-plasticity training in order to maintain the effect in the long term.

<u>*Preliminary methods and results*</u>

A schematic illustrating the visual search training task is shown in Figure 1. The task was practised on a notebook computer placed 30 cm from the patients’ eyes, covering a total visual field of 35 deg × 47.7 deg. Custom software was used to generate a random array of stationary digits or letters, 0-9; A-Z; size adjusted to the individual patient’s need, distributed with equal probabilities on the blind and seeing parts of the VF. RP subjects had to find and move the mouse pointer over the predefined digit or letter, for example, digit “6” as shown in Figure 1. Upon passing over the digit, the program generated a beep, recorded the time it took the subject to find the single target (RT per target) and turned it into a red symbol, which provided positive feedback and prevented double search for the digit. Importantly, during training, guidance related to where the targets might be, was not provided to the patients, neither eye movements were recorded. Thus the training was *static* and restricted to serial and random *search task* to possible locations in the non-seeing visual field. After all digits were found, the time between the initial screen onset and last target was recorded (RT per screen). The screen was then automatically cleared, and the patient started the next trial by clicking a button centered on the screen, which ensured initial central fixation for the next trial. Position and RTs for all digits found for every screen were stored in a database for each daily session. Initially, patients came to our laboratory and got instructed of how to perform the training. The subjects had then to train at home, using our laboratory’s notebook computers that ensure standard training conditions. EST was performed twice per day for 30 minutes, 5 days a week, for 6 weeks.

The effect of training that transferred to functional benefits in real life, was assessed by percent preferred walking speed, a measure of relative slowing, and eye movement behaviour before and after training in light (photopic vision at 65 cd/$m^2$) and dim (close to mesopic vision at 5 cd/$m^2$) test conditions. The percent preferred walking speed (PPWS) is a measure of relative slowing and was calculated according to the following formula: PPWS = (WS / PWS) * 100. Walking speed was measured in meters/second, while the mobile eye tracker (Tobii glasses) was used to record eye positions while walking in the standardized mobility course. The mobility course was a 68 m long and 1.3 m wide hallway containing 30 stationary obstacles that were arranged in pseudo-random positions to ensure an approximately equal amount of obstacles on the left and right side. While subjects walked the course, we also counted the number of errors, which were defined as the number of contacts with obstacles.

**Eye-movement analysis** We calculated the total number of fixations and average fixation durations for each subject and each condition (saccade or reading training). We categorized the fixations made by the patients outside their intact visual field (saccades into blind areas). The detection was performed using a velocity-based algorithm for saccade detection proposed by Engbert and Kliegl (36). The algorithm labels those eye movement episodes as saccades that show a velocity exceeding a certain threshold. Anything between two saccades is considered a fixation. A similar algorithm, based on a velocity threshold detecting method, was shown to adequately detect saccades in data collected by a mobile eye tracking device (37, 38).

Walking speed and eye movements data were collected at three times: pre- (T1) and post- (T2) training, as well as at 6 weeks after the training (follow-up at T3).

**Percentage of preferred walking speed** Average PPWS for the healthy control subjects was 92%, which was higher than for any of the RP subjects groups: saccade training (59%), and reading training (55%). Pairwise t-tests showed that PPWS at T1 and T2 did not change in either the healthy (non-training) or reading control groups ($p > 0.05$ and $p > 0.05$, respectively). Figure 2 shows the PPWS for the different training groups (saccade, reading and waiting list) before and after training (T1 and T2) in the different lighting conditions. Only in the light condition, walking speed in the saccade training group improved more after training than in the reading group.

**Number of errors** The average number of errors in pre- and post-training made by the saccade and reading training patients was significantly higher ($p < .001$.) in the dim (number of errors n = 12) than in the light condition (number of errors n = 3). The number of errors in the pre- and post- training phase for the different groups did not change significantly: The saccade training patients made on average 7 errors before and 6 errors after training ($p = 0.1$), while the reading training group made on average 8 errors before and after training.

**Number of fixations and fixation duration** Table 1 lists the frequencies of the fixations per minute, the average fixation duration, and the number of fixations beyond the VF in RP patients and the healthy control group. Only the subjects who were in the saccade training group made shorter fixations after training in the light condition ($F = 4.119$, $p = 0.03$). Table 1 also shows that before and after training, the RP subjects in the reading and saccade training groups, on average, directed more than 30 percent of their saccades to a region outside of the intact VF.

| Light condition | | | | Dim condition | | | |
|---|---|---|---|---|---|---|---|
| **Group** | **FF (T1/T2)** | **FD (T1/T2)** | **Nout (T1/T2)** | **Group** | **FF (T1/T2)** | **FD (T1/T2)** | **Nout (T1/T2)** |
| saccade | 39.8/47.9 | 2.8/1.1 | 16.3/19.7 | saccade | 31/32.9 | 3.5/ 1.77 | 8.6/9.2 |
| reading | 47.01/60 | 1.14/1.95 | 20.0/21.4 | reading | 55.8/28.2 | 1.9/2.8 | 17.5/11.4 |
| healthy control | 31.9/20.1 | 2.1/ 3.05 | | healthy control | 37.3/17.2 | 3.12/ 3.7 | |

*Table 1. Fixation durations (FD, in s), frequencies of fixations (FF, in fixations per minute) and number of fixations outside the seeing VFs (N out , in fixations per minute) of patients with RP in the reading and saccade training groups. Performance in the light and dim experimental conditions is shown before (T1) and after (T2) training.*

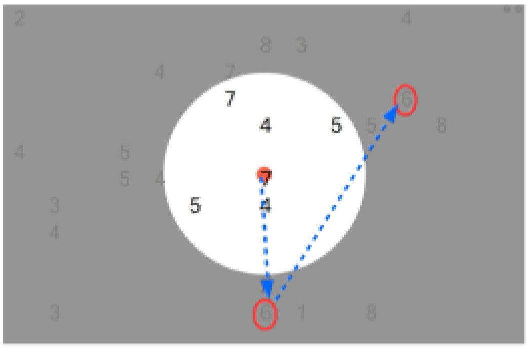

***Figure 1**. A schematic illustrating the visual search training task in our preliminary experiment. In this screen shot, the targets (number 6) are embedded among a random array of digits as distracters. It was possible that multiple stationary targets simultaneously appeared on the screen. After initial fixation of the central red dot, subjects were required to perform saccades (indicated by the blue arrows) outside their seeing visual field (gray area) to find all the targets (in the schematic surrounded by the red ovals). Each screen was presented until all targets were detected. The time-out period between the different screens was determined by the subjects.*

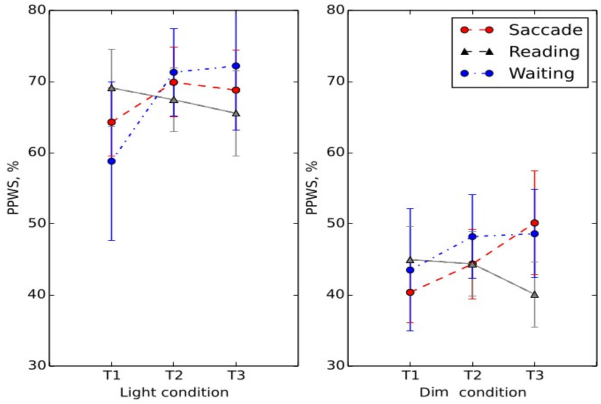


***Figure 2**. Separate plots present different test conditions. Performance was evaluated in light (left plot) dim (right plot) conditions for the saccade (red circles), reading (gray triangles) and waiting list (blue circles) training groups. The average percentage of preferred walking speed (PPWS) is plotted on the ordinate as function of the different times relative to the training: pre (T1)-, post (T2)- and follow-up (T3). Higher PPWS values represent better walking performance. Error bars represent +/-1 standard error of the mean.*

<u>*Summary of preliminary results*</u>

Before and after training, patients' performance was tested in a randomized and controlled trial using outcome variables such as, walking speed and time, number of collision with obstacles, number of saccades and fixation duration, parameters that are relevant to everyday life. After training, the walking time was found to be shorter, indicated by increase in relative walking speed, while number of collision with obstacles did not change. At the same time, after the training, the average fixation duration of the patients was significantly shorter. The improvement in patient relative walking speed due to shorter fixation durations indicates that training was beneficial for RP patients in this aspect. These findings further demonstrate that neural plasticity, via sustained, top-down attention can be consciously controlled and trained independently of the visual input.

<u>*Outstanding questions*</u>

Although, previous work also demonstrated some effect of EST, the impact on activities of daily living was unclear (39). In this study (39) part of the tests used to assess the effect of training tended to be very similar to the exercises practised during training. Conclusion from this study is that effect of training is maximal when tested on a similar task. In our preliminary experiment, however, we incorporated a standardized mobility-related test and demonstrated evidence for transfer of EST effect to activities of daily life beyond the specific task that was trained. These are encouraging findings that demonstrate the plasticity of the human visual system and its ability to benefit from such a training where conditions are static and lack the dynamics of situations in real life. <u>**The first outstanding questions,**</u> however, is that after EST, not all aspects of daily life improved with the training: number of collisions with obstacles and possibly with other people in the streets did not change. This suggests that further, improved training strategies might be required to ultimately optimize navigation of RP patients in daily life. We hypothesize that to maximize transfer of training effect in real life, exercise during training should resemble closely typical daily tasks. This could currently be achieved via virtual reality (VR) applications, with the added benefit that the patients could exercise to walk and deal with different situations without being exposed physically to the hazards of daily life.

Further implications about the limitations of the EST are related to information processing in human motion perception. To achieve a coherent perception, the visual system of the observer engaged in the activity must entail global integration of different motion signals (optic flow, object of biological motion) over space and time (40). This seems not to be a trivial task since optic flow, object and biological motion perception require different visual information processing and also different brain regions have been found to response selectively to the different signals. It is suggested that cortical area MST neurons are specialized to detect optic flow motion and have very large receptive fields, which respond selectively to complex optical flow fields, such as expansion, contraction and rotation. Neurons in area STS respond selectively to biological motion, while neurons in visual area MT are selective to object motion (for a review see 41). In contrast, our current training procedure is static and serial. In other words, patients are trained to find, by eye movements, non-moving, stationary targets on a computer screen. In this case the visual motion caused on the retina by the eye movements is not perceived due to saccadic suppression and the scene appear stable to the subjects (42). However, in our test phase, where effectiveness of the training is tested, conditions are different: Patients are required to walk and avoid obstacles. Although in this condition the environment is completely stationary, apparent motions of objects, surfaces and edges in the scene will be perceived due to the optic flow induced by the observer's own movement (43). This condition still lacks the full dynamics of more complicated daily life situations, where RP patients face extreme difficulties (4). For instance, to safely cross a busy intersection, static and moving obstacles, such as cars and other pedestrians, must be detected and tracked. In such a dynamic condition, object and biological motion together with optic flow perception play important role in order to track in parallel multiple targets, moving randomly over a range of retinal locations.

*Multi tracking and attention*

The human ability to track multiple targets in parallel is demonstrated in daily life, video games, sports, as well as in laboratory tracking tasks where subjects were shown to be able to track simultaneously 4 or more target on a computer screen (44-46). In several experiments it was shown that the ability to track simultaneously several targets is due to allocating attention in parallel across multiple independent non-contiguous loci in the visual field (44-46). This multi-focal "spread" of the attention across the independent loci in the visual field is in contrast to the classical notion of single loci of selection, where only one location at time may be attended to. Although the exact mechanisms by which attention keeps track of the simultaneously moving targets are still not well understood, it was found that spatial resolution of multi-focal attention is rather coarse and tracking performance degraded at small viewing angles (47). More recently it was demonstrated that eye fixations during multiple object attention were compatible with the notion of simultaneous multi-focal attention at discrete spatial loci and incompatible with serial selection (48). In other words, to estimate and keep track of multiple object positions, direct eye movements towards the moving objects were not performed by the healthy human visual system during tracking. Rather, attention was spread simultaneously across the whole natural field of view, spanning 210 deg horizontally and 150 deg vertically (49), to encompass all points of interest (47, 48). On the other hand, persons with RP have impaired mobility when visual field widths are with size of 30 degrees or less. This limited field of view in RP, as compared to natural vision, implies not only that RP patients are unable to get the global framework, due to lack of visual input from the periphery, but also that simultaneous multi tracking at retinal loci that fall outside functional areas may not be possible. This brings us to the challenging **second outstanding question**, which is to develop an efficient training that allows to compensate for the loss of simultaneous tracking of multiple moving targets in areas falling outside the limited functional VFs in tunnel vision. It was suggested that eye movements play an important role in global scene content perception, where the active contents of visual short-term memory (VSTM) are integrated with subsequently perceived information (50). In this study (50), it was demonstrated that in different static or dynamic conditions, healthy subjects used distinct encoding strategies: Information was integrated covertly, via multi-focal attention or overtly via successive eye saccades. Therefore, we hypothesize that multi tracking in RP may be supported by alternative mechanisms, for example sustained covert attention and rapid serial scanning of possible loci of interest to retinal areas that lack visual input.

#### 1.1 Project-related publications

##### 1.1.1 Articles published by outlets with scientific quality assurance, book publications, and works accepted for publication but not yet published.

Barraza-Bernal M, **Ivanov** IV, Nill S, Rifai K, Trauzettel-Klosinski S and Wahl S. (in press, Vision Res DOI:10.1016/j.visres.2017.07.009) "*Can positions in the visual field with high attentional capabilities be good candidates for a new preferred retinal locus*?"

**Ivanov** IV, Mackeben M, Vollmer A, Martus P, Nguyen NX, Trauzettel-Klosinski S. (2016) *"Eye Movement Training and Suggested Gaze Strategies in Tunnel Vision - A Randomized and Controlled Pilot Study."* PLoS One. 2016 Jun 28;11(6):e0157825

Trauzettel-Klosinski S, **Ivanov** IV, Damm I and Reinhard J (2014) "*Eye movements during saccadic and fixation tasks in patients with homonymous hemianopia.*", J Neuroophthalmol. 34(4):354-61

**Ivanov** IV, Leitritz MA, Norrenberg LA, Dynowski M, Ueffing M and Dietter J (2016) "*Human-vision motivated algorithm, allows consistent retinal vessel classification based on local color contrast for advancing general diagnostic exams.*" Invest Ophthalmol Vis Sci. 57:731–738

**Ivanov** IV, Kramer DJ and Mullen KT (ePub 2013) "*The role of the foreshortening cue in the perception of 3D object slant*" Vision Res. 94:41-50

##### 1.1.2 Patents

###### 1.1.2.1 Pending

"A new class of optotypes to asses habitual refractive errors, the contrast sensitivity and the neural transfer function of the eye under consideration of the global visual integration of the visual system", European Patent Office reference number 16002585.4.

### 2 Objectives and work programme

#### 2.1 Anticipated total duration of the project

The anticipated total duration of the project is four years. DFG funds are requested for two years. The preliminary results for this study were supported by: Kniese Foundation; Funke Foundation; Stiftung Auge; Pro Retina to Prof. Dr. Trauzettel-Klosinski and AKF-Programm, University Eye Hospital, University of Tuebingen, Grant Nr. 296-0-0 to Dr. Iliya V. Ivanov

#### 2.2 Objectives

**Main hypothesis** As outlined in the *State of the art and preliminary work* section, to safely navigate in challenging environments, where parallel tracking of multiple moving objects may be vital, is the hardest problem that RP patients face in their daily life. We hypothesized that via neural-plasticity training, visual performance in RP can be restored. We choose indicators of visual performance restoration related to daily life: percent preferred walking speed and number of contacts with obstacles.

**Objective 1** To develop and validate a neural-plasticity training method able to compensate for the limited visual performance and daily life quality in patients with RP and to improve the awareness about the size of the visual field defect. The efficacy of the new training is quantitatively evaluated by testing patients' visual function in a visual search task and performance in a standardised daily life mobility task before and after training.

**Objective 2** To maximize the amount of restoration in visual function and improvement in daily life performance of RP patients. In our preliminary results the training relied on the natural abilities of the patients to develop successful compensatory eye-movement strategies and the task was to find only stationary targets. Although visual performance improved, as indicated by the shorter fixation durations and walking times after training, number of collisions with obstacles, an important daily life parameter, did not change with training. Therefore, the main goal here is to minimize the number of collisions to a level that would allow safe and independent navigation of the RP patients in unfamiliar environments.

***How we are going to achieve the objectives?***

**Objective 1, first results** In our preliminary work we demonstrated that neural-plasticity training to allocate sustained attention to areas in the non-seeing VF resulted in significant improvement in visual search and mobility performance in daily life. We implemented the training as a task that rely on the natural abilities of the patients to develop successful compensatory eye-movement strategies.

**Objective 2** A new training method will be devised and validated. In a first step, the new training will instruct the patients to perform in a systematic scanning pattern (SSP) fast compensatory gaze movements, in order to create a global framework of the scene and identify possible obstacles (targets). In a next step, dynamic scanning tracking (DST) of possible targets will be practised, like in the switching model of multi tracking proposed in (45, 51, 52), which requires only one focus of attention that must cycle rapidly through the targets, indexing their locations and returning to each before it moves too far away. As attention will revisit each remembered object location in the SSP, the nearest item will be taken as the new position of the target, and that location would be stored for the next cycle. The rational here is that by applying the DST, the subjects would be able to see the objects repeatedly and thus it will be easier to track them (53). Our aim here is to employ SSP and DST as a blueprint for a new method that will improve visual performance in patients who have functional VFs less than 30 deg, a size too small to allow for instantaneous perception and to support simultaneous multi tracking within the global scene framework.

This new method builds on our current EST in several important aspects:

*1. Systematic and dynamic.* The new training method will instruct patients in a systematic manner to perform SSP and DST via overt and covert shifts of sustained attention in order to create a global environmental representation and to detect, as well as to track, possible targets within a dynamic framework. For detailed description of this training implementation, please refer to the *Methods* section on pp. 11-13.

*2. Pervasive and adaptive assessment of performance.* Real time gaze control during training and practising will be performed via the use of mobile eye tracking and virtual reality (VR) with embedded eye tracking technologies. In this way, performance and progress will be monitored and assessed in real time without any feedback required by the patients. Further, difficulty of the practising task will be adaptively adjusted as function of subjects' current performance level. Thus, training time will be optimized for each patient individually, engagement and activity enjoyment will be increased and thereby clinical outcome will improve.

*3. Practising in a virtual reality context to stimulate transfer to daily life.* To create a realistic, interacting and immersive context of practising environment, technologies such as VR and head mounted displays (HMD) will be employed. We will implement DST in a virtual reality game context, where dynamic simulation in VR rehabilitation may reduce the difference of movement strategies in the virtual and real world. Here, the reduced difference between eye scanning patterns, will be the key factor to enhance the transferability of virtual skills to the real world (54-56). Further, the use of VR and HMD will allow the patients to develop natural compensatory strategies involving eye, head and body movements, whereas contemporary EST method relies on eye movements only. In EST, visual field coverage is smaller, as compared to VR training, where angles up to 360 deg are possible.

#### 2.3 Work programme incl. proposed research methods

**Ethics** The preliminary experiment of this study was approved by the ethics committee of the University of Tübingen Medical Faculty, and informed written consent was obtained from all participants in the preliminary experiments. The research adhered to the tenets of the Declaration of Helsinki. Before the new experiments commence, a separate approval by the ethics committee of the University of Tübingen Medical Faculty will be requested.

**Subjects** A cohort of 30 patients with documented retinitis pigmentosa will be recruited for the study from the clinical data base of the Low Vision Clinic at the University of Tuebingen. An age-matched cohort of healthy subjects (n = 15) will also be recruited as a control group. Inclusion criteria for the patients are no other eye diseases, visual acuity (VA) higher than 0.1 dec (1.0 logMAR) and maximum visual field (VF) size of less than 30 deg of visual angle. Patients at different stages of the RP development will be recruited. To avoid any confound of the results due to possible differences in adaptation, patients will be assigned randomly to the different training groups (see *Study design*) and training practise phase will be adaptive (see *2.2.2. DST practise*). All healthy controls will have normal or corrected to normal vision. Patients with coexisting eye movement pathologies or cognitive impairments will not be recruited in the study.

**Study design** In a waiting list control group design, subjects will be randomly assigned to experimental training (SSP and DST group, n = 15) or control training (EST group, n = 15). For ethical reasons and to ensure that all patients will have a chance to benefit from the study, the patients, who will first be assigned to the control training, afterwards will also undergo experimental training (waiting list group). Additionally, data from 15 healthy, normally-sighted observers will be included as a non-training control group (healthy control group). This group will not receive training but will undergo the same visual function and performance tests as the patient groups over the same period of time. Thus, the healthy control group will reveal the degree of performance change that would occur without training. The disclaimer here is that it would only reveal this for healthy controls. In all RP patients, before (T1) and after (T2) training (SSP, DST or EST) and six weeks at follow-up (T3) visual performance of patients on a multi-focal tracking task and two types of mobility courses (virtual and real-life) will be assessed. On the same tests, performance of the healthy controls will be assessed at T1, T2 and T3. Performance of the two patient groups at T1 will be compared to performance of the healthy control group and changes between T1 and T2 in the experimental training group will be compared to changes in the control and waiting list groups.

**Methods**

**Visual function tests** The following main standard visual function tests will be performed on all patients before and after taking part in the training: visual acuity, contrast sensitivity, monocular visual field perimetry with continuous control over stability of fixation. For the healthy control group, only visual acuity will be controlled.

**Available training**

**EST** Exploratory saccadic task (EST) was implemented to train sustained attention and evaluate effect of training in a mobility task. Detailed explanation of the EST method was given above in the *State of the art and Preliminary* work section.

**Training to be developed**

**SSP training** The aim of this training is to teach patients with RP of fast systematic gaze movements, in order to create a global framework of the scene and identify possible obstacles (targets). Patients will be taught a systematic scanning pattern (SSP) consisting of a sequence of small and large saccades, as well as head movements along the four cardinal directions of the compass. First, a forward scanning strategy along the west (*W*) cardinal direction will be executed (see Figure 3): it starts with one small saccade (indicated by the arrow to *s1*) ending in the periphery of the intact concentric field (blue dotted circle with radius *R*), followed by a large saccade to *s2* and a possible head movement (*h2*) towards the blind field. The size (*2R*) of the larger sacade would be twice the radius of the intact visual field, so there is no overlap between the VF coverage of the two consecutive saccades, unlike in the small saccade case (shaded area in Figure 3). In this way, the role of the small saccade, which always falls inside the seeing VF, is to prepare the execution of the larger saccade that is in the same direction but falls into the non-seeing VF. As in normal scanning behaviour, should the size of the required larger saccade (*s1* to *s2*) exceeds 44 deg, the largest saccade that can be done without head movement, subjects would be required to perform a head movement (*h2*) to stabilize the gaze in the centre of the VR scene (outermost grey rectangle). The systematic sequence *s1 + s2 + h2* will be executed *n*- times until the total size of the VF coveredin this direction is close to 100 deg (see Figure 3). Then the subjects will be required to execute a free backward sequence, for example *s1 + s1 + s2 + h2 + s2 + h2*, from the end-point of their forward sequence ($O_1$), in order to return to the initial fixation *O* (Figure 3). The same duo of forward and backward sequences will then be executed along the east (*E*) cardinal direction. The SSP strategy will continue further with forward and backward sequences along the north (*N*) and south (*S*) directions, where sequences are narrowed to 75 deg coverage. In this way patients will be trained to compensate for their loss of field of view and dynamically expand it, along the compass cardinal directions, to approximately the size of a normal field of view: 2 * 100EW = 200 deg horizontally and 2 * 75NS = 150 vertically. Note that in VR angles up to 360 degrees could be covered. However, we narrow our training to the filed of view naturally covered (200 x 150 degs). The SSP will be implemented in a virtual reality HMD environment, rendered by FOVE 0 (FOVE Inc., Figure 5), a virtual reality googles featuring low latency stereo eye tracking. Note, that since at the time of writing FOVE 0 is available as pre-order only, an alternative device is also considered (see p. 17, in the *Requested modules* section). During forward and backward sequence execution, small circular targets (black dots in Figure 3) will instruct the patients to perform *s1* and *s2* saccades. Current eye position will be analysed in real time loop via the FOVE 0 eye tracker and continuous auditory cues (*A*) will guide the adjustment of eye movements ($s_1$ and $s_2$, Figure 3) until they precisely (within 1 deg) reach the targets s2 (57). Here, the use of continuous auditory cues (sonification), in contrast to simple auditory feedback or sound alarms, will provide feedback generated concurrently to the eye position real-time analysis and thus the patients will be informed on the precision of their oculomotor behaviour. The patients will be instructed to adapt their fixations according to the sonification, generated by the hosting computer. For instance, four distinct pitches of sound would indicate the need of left- and right- or up- and down- ward correction of the fixation along the horizontal and vertical meridians respectively. Additionally, the loudness of the sound could also be manipulated as function of distance between current eye and target positions. In this way smooth and continuous sonification of the dynamic range of the eye motion would be achieved. Patients will learn to generate this systematic scanning pattern endogenously on an anticipatory basis, training half an hour each day for six weeks (21 hours in total). In a next step, they will practise to adjust the speed of repetition of this scanning pattern to environmental demands and to the speed of walking (see DST practise below).

**DST practise (VR rehabilitation)** Dynamic simulation of the DST practising task will be implemented using state-of-the-art game engine by Unity Technologies rendered on FOVE 0. Unity engine provides free licenses for educational, non-commercial applications where behaviour of virtual objects and their responses to external force and torque are simulated in a physically realistic manner. A virtual reality model of a city will be created with objects that look and act like real. Patients will be required to practise the systematic saccadic patterns instructed in SSP training phase in a dynamic setting: to move within the VR city community and avoid collisions with obstacles, like other pedestrians, that could be encountered in real-life in different situations. (As an outlook, but outside the scope of this proposal, a motion tracking system may be used to enable patients to really move and at the same time benefit from the safety of the VR.) VR provides control over obstacle positioning, which could be standardized, and participants will have to make their way through the course in order to avoid touching the obstacles. The mobility situations will adaptively build up from quiet, structured and familiar surroundings to busier and more complex and unfamiliar surroundings (imaginary city with parks, buildings etc.), depending on the current progress of the patient. The patients' progress in the practise session will be assessed in real-time via checking the state of physics simulation model and gaze position: to do so, number of collisions and gaze fixations at obstacles will be calculated and analysed. Then, like in a slow motion reply, the dynamics and the physics fidelity of the simulation will be adjusted individually to the current performance level. In this way, the patients will learn to repeatedly re-apply SSP and thus to cycle rapidly through the targets, will index their locations and will return to each before it will move too far away. This adaptive phase of the training is ultimately aimed to make the systematic scanning pattern default, automated activity embedded in every mobility situation encountered by the patient in daily living. The patients will be required to practise in 15 min blocks, 2-3 times daily, during the six week of SSP training. To maintain possible effect of SSP and DST training in the long run, patients will be required to keep practising 2-3 times weekly.

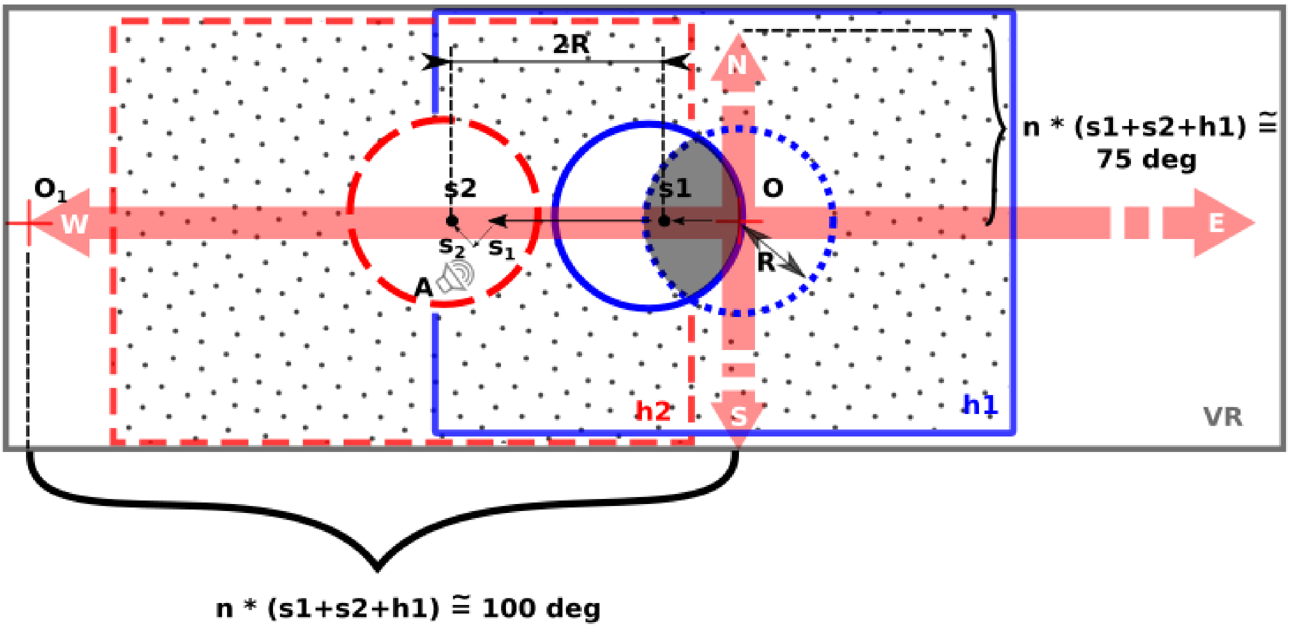


***Figure 3.*** *A schematic of the systematic scanning pattern (SSP) training. The four cardinal directions are represented by the salmon arrows. Red cross at O indicates initial eye and head (h1) fixation in the VR (outermost gray rectangle VR). SSP starts with one small saccade (indicated by the arrow to s1) ending in the periphery of the intact concentric field (blue dotted circle with radius R), followed by a large saccade to s2 and a possible head movement (h2) towards the blind field (doted areas). The size (2R) of the larger saccade is twice the radius of the intact visual field, so there is no overlap between the VF coverage of the two consecutive saccades, unlike in the small saccade case (shaded area). The systematic sequence s1 + s2 + h2 will be executed n- times until the total size of the VF covered is close to 100 deg along the E and W directions and 75 deg along the N and S directons. Forward to $O_1$ and backward to O sequences will be instructed. Real time sonification (A), generated by the hosting computer, will guide the adjustment of the eye movements ($s_1$ and $s_2$) until they reach precisely the targets s2, falling in the blind VF of the patients.*

##### Performance indicators

**Visual tracking task** will be implemented as a multi-focal tracking test. We will perform this test since, to the best of our knowledge, it is not known whether multi-focal tracking is spared in RP like in healthy subjects. Figure 4 shows the stimuli and the presentation sequence that will be used. Observers will view a number of squares, a subset of which will be briefly highlighted to indicate the targets to be tracked. A randomly chosen subset from one to five of the total field of twelve objects will be designated as targets. Each object will subtend a visual angle of 0.4 deg and will move with a velocity and direction changed at random every few hundred milliseconds. The velocities of the squares will range from 1.25 to 9.4 deg/s. The directions will be chosen from among 8 equal divisions of the compass. The random motion of the objects will be restricted, so they can not be closer than 0.75 deg apart: In this way the continuity of the objects identity would never be ambiguous, as it would be if they were allowed to collide. At the end of each trial, the squares will stop moving and observers will be asked to select all the targets. The sequence will be presented via a HMD, e.g. FOVE 0, and targets will be selected by the observer via positioning and maintaining a small dot presented in the centre of the goggles for 1 second at the target. Feedback will be provided, such that the successfully identified targets will be marked with the star symbol, as indicated in Figure 4. Accuracy, reaction times and eye movements will be recorded and analysed before and after training.

**Mobility course, static indoors setting** The same course used in the preliminary EST experiment would be employed here. Importantly, in contrast with the preliminary experiment, a device with higher sampling rate for the eye tracking will be used to allow for complete analysis of related parameters of the eye-movement data collected during walking. The latest version of the SMI mobile glasses (SMI-ETG 2W), sampling at 120 Hz, will be used for the eye-movement data collection (58). For details of the mobility course, please refer to pp. 4 and 5, in the *Preliminary methods and results* section. To assess any effect of training that transfers to functional benefits in real-life, percent preferred walking speed, number of contact with obstacles and eye movement behaviour before and after training will be analysed. Safe and efficient orientation and mobility is a standard test to assess visual performance of patients with low vision (59)

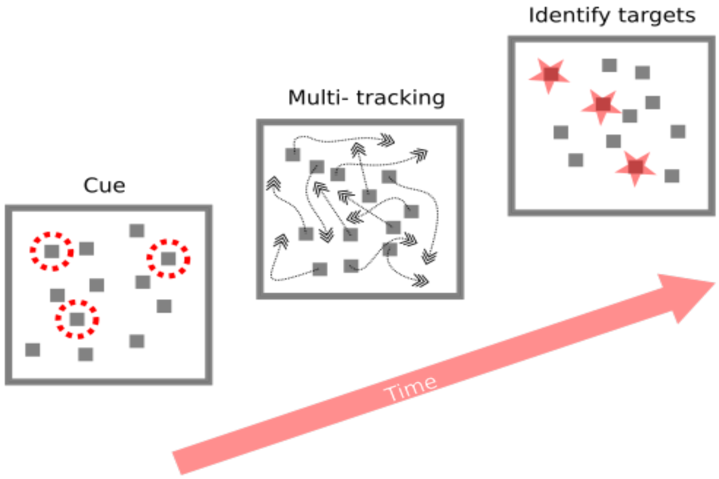


***Figure 4***. *An example of the multi-focal attention test. The target squares are highlight by the surrounding red doted circles. After a little while, highlighted circles disappear and all squares start moving. At the end of the trial movement stops and the participant is asked to select the target squares.*

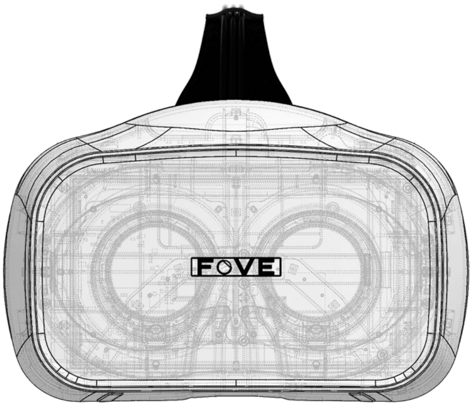


***Figure 5.*** *FOVE 0,(FOVE Inc.), a virtual reality googles featuring low latency stereo eye tracking.*

**Mobility course, VR real-life simulation** Dynamic simulation of the virtual environment will be implemented using state-of-the-art game engine by Unity Technologies rendered on FOVE 0. A virtual reality model of a city will be created with objects that look and act like real. The task performed by the subjects will be divided in two parts: 1) Familiarize with the environment and apply SSP to collect the global picture, time given is as long as the subjects want; 2) Way- finding task, to make their way, for example, to the nearest bank office and claim their virtual reward.

In the first part, participants will be given a training session to familiarize themselves with the environment. They will perform a free eye and head (gaze) movements during their navigation within an imaginary city and will receive no instructions other than to walk normally, avoid obstacles and to collect as much as possible knowledge about the environments (indoors and outdoors) and memorize navigation landmarks. The virtual reality will include segments from unfamiliar indoor (stairs, ramps, doors, elevators etc.) and outdoor (including traffic, pedestrian crossings, and naturally occurring obstacles) environments and city streets.

In the second part, after the training session, subjects will be asked to perform the actual way-finding task. They will be instructed to make their way to the nearest bank office and claim their virtual reward. All participants will start from the same position and no guidance will be provided. The participants main goal will be to navigate safely through the course, where contact with obstacles will result in penalty. For motivation, the amount of virtual reward will be inversely dependent on the number of contacts with obstacles and time to perform the task. Unlike in the DST practising session, there will be no slowing of the VR simulation dynamics and the physics fidelity, and thus interactions in this testing phase would be as in real-life. The effect of training that transfers to functional benefits in real life will be assessed by analysing percent preferred walking speed, number of contact with obstacles and eye movement behaviour (FOVE eye tracking) before and after training.

**Questionnaire** Quality of life of the patients will be assessed by a standardised German version of NEI-VFQ 25 questionnaire (4) before and after training, as well as at follow-up. The questionnaire includes the following categories with one to six items each: general health; general eyesight; eye pain; near vision; distance vision; vision specific impact on social functioning; vision specific impact on mental condition; vision specific effects on social role exercise; vision specific effects on dependence on others; driving; colour vision; peripheral vision.

**Assistance needed outside own group/institute**

In the own group expertise exists on testing multi-focal attention in RP patients, evaluating efficacy of the new training methods (Dr. Iliya V. Ivanov), as well as implementing virtual reality applications (Dr. Katharina Rifai). Expertise in conducting behavioural experiments with patients (Dr. Iliya V. Ivanov, PI) is available at the own Lab. Complementary clinical expertise and patient recruiting assistance are provided via our collaboration with the Lab of Prof. Dr. med. Dr.h.c.mult. Zrenner, the Vision Rehabilitation Research Unit, University of Tuebingen (Prof. Dr. med. Trauzettel-Klosinski) and the Low Vision Clinic (Prof. Dr. med. Nhgung), University of Tuebingen.

**Schedule for the planned experiments**

Time envisaged to complete the proposed work is 24 months. For a schedule detailing the steps outlined above during the proposed funding period, please refer to Table 2, Time frame.

#### 2.4 Data handling

Local data base storage will be created to store all data produced within the proposed research project, including the definition of metadata. The database will be securely connected with the local existing clinical data repositories.

#### 2.6 Descriptions of proposed investigations involving experiments on humans, human materials or animals

This is a non-interventional, observation study, similar to a computer game, on patients with retinitis pigmentosa and involves experiments on healthy observers. Inclusion criteria for the experiments are normal or corrected to normal vision for the healthy observers and documented retinitis pigmentosa for the patients with no restrictions on age and sex, while care will be taken to ensure similar average age for patient and control groups. No potential risks for the patients and healthy control observers are known. Information about the research and possible risks to the possible participants were/will be conveyed through advertisements, recruitment letters, pre-screening phone calls, study description sheets as well as written informed consent documents and discussions. The written informed consent documents and discussions were/will be conducted in 'lay' German and/or English language understandable to the subjects to contribute to their understanding of the research purpose.

### 4 Requested modules/funds
#### 4.1 Basic Module
##### 4.1.1 Funding for Staff

Requested for the project are funds to cover 1 post-doctoral (PD) student position at full-time (€136,800 in total) and 1 research assistant at 50 percent, (€53,100 in total) during the 24 months of the project.

###### 4.1.1.1 Postdoctoral researcher

The PD student will be responsible for development and implementation of the new training methods, the data processing and statistical analysis, including selecting the appropriate analysis procedure, setting up data for analysis, judging validity and reliability of data, designing and writing programs to perform analyses, and producing written reports on the result of the analyses. S/he will establish the daily priorities and recommend patient visit schedules. S/he will be actively involved in the presentation of project results to internal and external parties and engaged in writing of project results publications and reports. Directly supervise the research assistants of the project. Required qualifications are doctorate in vision science or closely related field and experience in programming VR. Good organizational and project management skills, as well as written and oral communication skills.

###### 4.1.1.1 Other research assistant

One research assistance will be recruited to assist the PI and the PD. Main responsibilities will be to conduct literature reviews, collect experimental data, recruit and interview the patients with the questionnaire, maintain accurate records of interviews, safeguarding the confidentiality of subjects, as necessary and summarize the interviews. S/he will perform and/or assist wit the assessment of patients’ visual function. Further duties are to provide ready access to all experimental data for the PI and PD and respond to project related emails.

##### 4.1.2 Direct Project Costs

The total amount of the direct project costs is €18,870

###### 4.1.2.1 **Equipment up to €10,000, Software and Consumables** (approx. €9,970 requested)

Requested equipment is for the VR-HMD training methods: FOVE 0 and training station notebooks with state-of-the-art graphics capable to support demanding VR rendering and real-time eye tracking calculations. The FOVE 0 eye-tracking VR googles (FOVE Inc., pre-order only, available for $599 at the time of writing), provides real-time streaming of eye and gaze data to allow for online gaze tracking in the VR. A gaming Dell notebook with graphics card that supports VR rendering is required. The cheapest option available at the time of writing is Dell Alienware 13 Gaming with Nvidia GTX 960M, from €1,479 at Dell.de. The total of 5 training stations (FOVE 0 and a Dell gaming notebook) at the cost of approx. €9,970 are requested for the purpose of the study. With 5 stations, that will be borrowed to the subjects to practice at home for the prescribed period of six weeks, the estimated time to complete the training only is approximately 9 months. Alternative and costlier VR implementation is possible via the combination of Oculus rift googles (€449 at the time of writing) and the Pupil Labs eye tracking add-on (€1,400 and €800 for the binocular and monocular eye tracking options, respectively).

###### 4.1.2.2 **Travel Expenses** (€5,000 requested)

Funds in the amount of €2,500/year are requested for presentation and dissemination of the results from the research to the scientific community. Presentations are envisioned at the annual Association for Research in Vision and Ophthalmology meeting and at the European Society for Low Vision and Rehabilitation meeting.

###### 4.1.2.3 **Other Costs** (€2,400 requested)

An amount of €2,400 is requested to compensate patients and subjects participating in the study. On average a “Pauschale” (covering travel costs) of €75 per patient is envisioned, as well as €10 hourly per healthy controls, locally recruited, so no travel costs are covered: 30 patients * €75 = €2,250; 15 Healthy controls * €10 = €150.

###### 4.1.2.4 **Project-related publication expenses** (€1,500 requested)

The maximum of €750/year is requested for the publication of the scientific result from the project.

#### 5 Project requirements

##### 5.1 Employment status information

Ivanov, Iliya V., wissenschaftliche Mitarbeiter, University Eye Hospital, University of Tuebingen, contract until end of 2018, further prolongation until the end of the proposed research is envisioned, see formal letter from Lab Director, Dr. Siegfried Wahl.

##### 5.3 Composition of the project group

Principal investigator (PI) Dr. Iliya V. Ivanov, MSc. Maria Barraza-Bernal, MSc. Alexander Leube, Dr. Katharina Rifai, Dr. Siegfried Wahl.

##### 5.4 Cooperation with other researchers

###### 5.4.1 Researchers with whom you have agreed to cooperate on this project

Prof. Dr. med. Dr.h.c.mult. Zrenner, Werner Reichardt Centrum für Integrative Neurowissenschaften, Dr. rer. nat. Siegfried Wahl, ZEISS Vision Science Lab, University of Tuebingen, Prof. Dr. med. Trauzettel-Klosinski, the Vision Rehabilitation Research Unit, University of Tuebingen, Prof. Dr. med. Nhgung, the Low Vision Clinic, University of Tübingen, Dr. med. Zobor, Institute of Ophthalmic Research, University of Tübingen.

###### 5.4.2 Researchers with whom you have collaborated scientifically within the past three years

Dr. rer. nat. Siegfried Wahl, ZEISS Vision Science Lab, University of Tuebingen, Prof. Dr. med. Trauzettel-Klosinski, the Vision Rehabilitation Research Unit, University of Tuebingen, Prof. Dr. med. Nhgung, the Low Vision Clinic, University of Tübingen

##### 5.5 Scientific equipment

The ZEISS Vision Science Lab is a working group for vision research and conducts rigorous research in order to spearhead the understanding of vision. Through science in seeing products and solutions for natural and enhanced vision are enabled in an academic environment that favours creativity for generating groundbreaking ideas and their translation into reality. The ZEISS Vision Science Lab investigates the complex interaction of light, the eye and the processing in the brain in different and dynamic situations. The goal of this research is to gain an understanding of the development of seeing and perception to develop new ways of providing natural, optimized vision. Another item on its agenda is to research into the development of vision and into pathological changes to perception in order to enable their diagnosis by using suitable measuring methods at an early stage. For these patients, this could result in personalized solutions for enhanced vision. The laboratory has considerable experience in various techniques in the fields of ophthalmology and optometry and is equipped with various ophthalmic devices. Additionally state of the art eye tracking and stereo systems are applied to learn about fixational eye movements and gaze strategies. The closed loop of using these instruments and understanding more about the eye and the processing in the brain will also lead to a continuous improvement or even to innovative new instruments for diagnosis or therapy in ophthalmology and optometry. The lab provides facilities to support research in physiological and clinical visual optics and in the field of visual neuroscience as well as neuro-computation.

**Table 2. Time frame of the project** ***"Improving visual performance in retinitis pigmentosa by new non-invasive personalized and adaptive training."***

| Project month | Objective | Activities | Expected completion (month) | Person(s) responsible |
|---|---|---|---|---|
| 0 | Demonstrate the effectiveness of explorative saccade training (EST) in daily life in tunnel vision | 1) Recruit a cohort of 25 patients with documented retinits pigmentosa<br>2) Conduct pre- post- and follow up assessments of visual performance, related to daily-life of RP subjects after applying EST neural-plasticity training for 30 min, daily for 6 weeks<br>3) Data analyzed and results published at Plos One<br>4) Apply for approval of the new methods by the ethics committee of the University of Tübingen Medical Faculty | Preliminary work, already completed | Principal investigator (PI) |
| 1 | Develop new SSP training and DST practise methods; implement visual multi-tracking task test | Code specification, implementation and extensive testing of the new training, practice and test methods: SSP training, DST practice and multi-tracking visual performance assessment test | 5 | Post-doctoral student (PD) and research assistant (RA) |
| 6 | Recruit the cohort of 30 RP patients and 15 healthy controls | 1) From the Low Vision Clinic database, screen, contact and recruit suitable patients to participate in the study, via telephone and/or email<br>2) Arrange for patients' travel to and from the Lab to get training instructions and notebooks; Most must travel long distances<br>3) Prepare and obtain informed written consent from all participants willing to take part in the experiments | 7 | PD and RA |
| 8 | To improve visual performance and mobility of patients with RP:<br>- Apply new SSP training and DST practise methods;<br>- Assess visual performance and function at pre-, post- and follow up training phases; | 1) Initial (pre-) assessment of visual function (standard visual acuity, contrast sensitivity tests, visual field perimetry etc.) and performance (multi-tracking test, mobility in VR and real environment and questionnaire) of all subjects<br>2) Patients (n= 30) randomly assign in different training groups (experimental and control) and start practising at home on 5 training stations for the prescribed period of six weeks each: approximately 9 months in training only<br>3) Collect, analyse and report at one scientific conference intermediate results<br>4) Post- training assessment of visual function (standard visual acuity, contrast sensitivity tests, visual field perimetry etc.) and performance (multi-tracking test, mobility in VR and real environment and questionnaire) of all subjects<br>5) Start preparing final results for publication<br>6) Final (follow-up) assessment of visual function (standard visual acuity, contrast sensitivity tests, visual field perimetry etc.) and performance (multi-tracking test, mobility in VR and real environment and questionnaire) of all subjects<br>7) Collect, analyse and report at one scientific conference final results<br>8) Finalize results of the new methodology and behavioural training for publications | 23 | PD and RA |
| 24 | | Prepare final project report to DFG | 24 | PI; PI advises and monitor project budget and execution at all times |
