## Supplementary material for "Influence of open-source virtual-reality based gaze training on navigation performance in Retinitis pigmentosa patients in a crossover randomized controlled trial": S1 Appendix

### Appendices

#### Appendix A - Scanpath Evaluation using Multimatch-Algorithm

As was described in the 'Scanpath' section, the patients' gaze movements during training were measured, and the similarity between this gaze movement and the suggested scanpath was calculated at run-time. To do so, the first step is to analyze the eye-tracking data captured by the VR device to determine saccades executed by the user. Saccades describe the rapid eye movements found in-between fixation points of the gaze. A saccade is detected when the gaze velocity surpasses $50°/s$ (based on Gibaldi et al. [30]) and when the direction of gaze movement does not change by more than 30°. Using a modified Multimatch-Algorithm [26] , sections of multiple saccades executed by the participant are compared to a saccade representation of the suggested Scanpath (Fig. 12) to calculate a similarity value based on how well the two saccade patterns match. This similarity value was displayed to participants after each trial of the Gaze Training, giving them a quantitative measure of how closely their gaze movements match the suggested Scanpath.

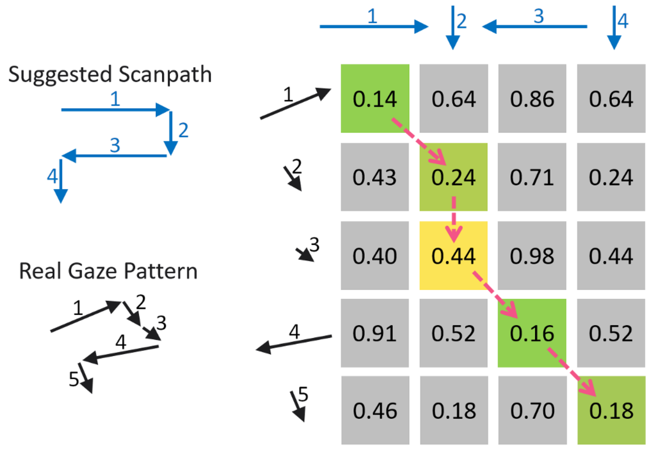


Figure 12: Visualization of a Multimatch Scanpath comparison between a gaze pattern displayed by a participant (Real Gaze Pattern) and an ideal representation of the suggested Scanpath. Each square displays the difference in angle and amplitude between the respective saccade vectors, with 0.0 meaning saccades are identical and 1.0 meaning saccades are complete opposites. Colorized squares indicate the "path of the least resistance" determined by the Multimatch-Algorithm, which describes the best match between the two patterns.

#### Appendix B - Patients' Fields of View

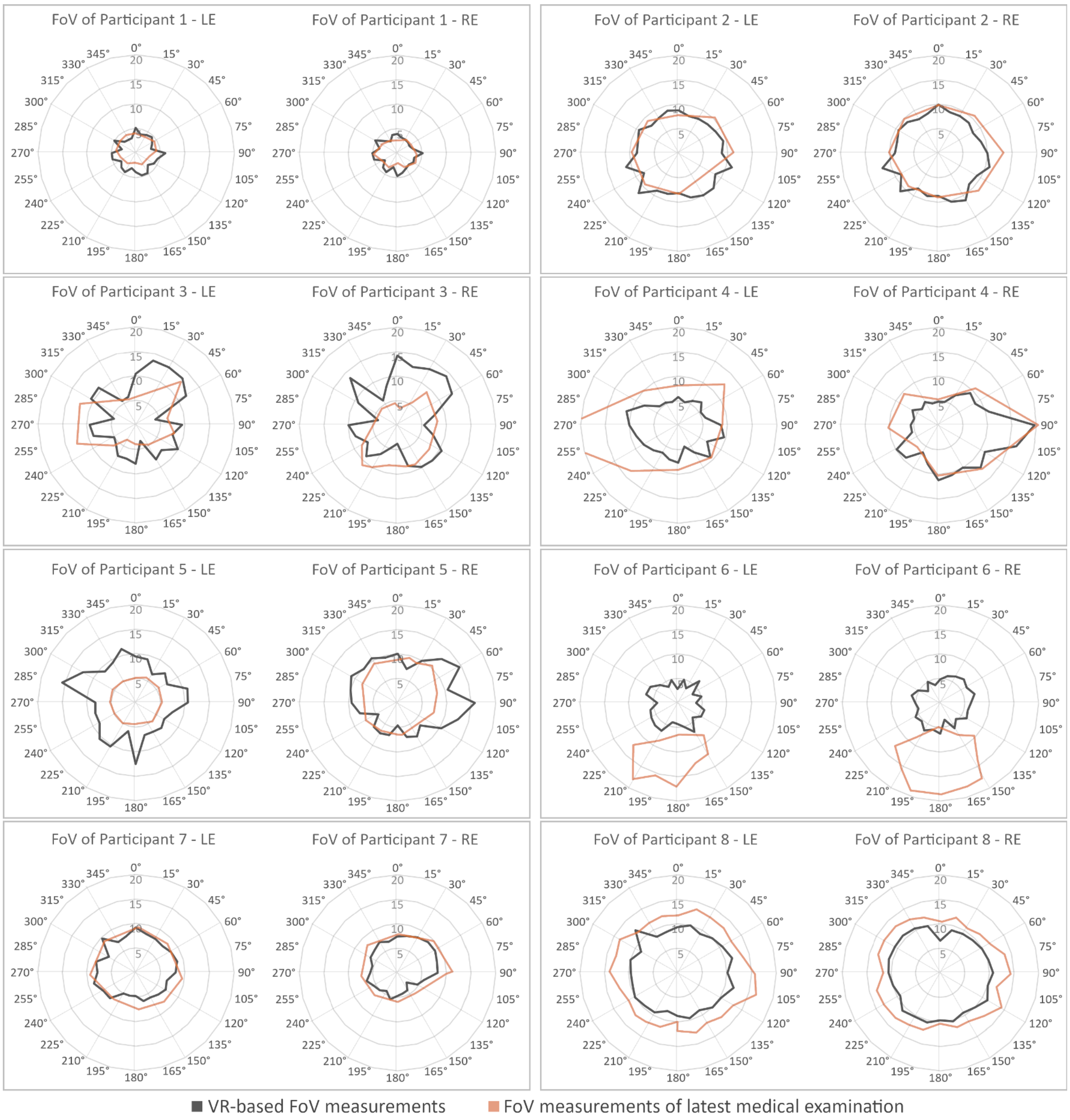


Figure 13: Visualization of the visual field dimensions of the eight participants who completed the study. Grey indicates the VF measured by the self-developed, VR-based kinetic perimetry tool as described in Chapter 3.2.2, orange indicates the participants' VF based on their most recent medical examination.

#### Appendix C - Statistical Models and QQ-plots

This section lists full details on the models used for the statistical analysis of the real-world obstacle course results, as well as the QQ-plots used to visualize normal distribution of results.

**Effect of Gaze Training (pre/post training condition) on trial duration**

Model Specification:

- Model: Linear Mixed Model (lme)
- Dependent Variable: log(TrialDuration)
- Fixed Effects: PrePostTrainingCondition
- Random Effects: $\sim 1 +$ Participant | Participant
- Model Fit Statistics: AIC = -285.7289, BIC = -263.1566, logLik = 148.8645

Results:

- Intercept: Estimate = 3.567655, SE = 0.09670653, t-value = 36.89157, $p - value <$ 0.001
- PrePostTrainingCondition: Estimate = -0.177829, SE = 0.01577808, t-value = -11.27062, $p - value < 0.001$

Effects:

- Participant (Intercept): StdDev = 2.717010e-01
- Participant: StdDev (Intercept) correlation = 0
- Residual: StdDev = 1.411235e-01

Data Samples:

- Number of Observations: 320
- Number of Groups: 8

**Effect of control phase (pre/post control condition) on trial duration**

Model Specification:

- Model: Linear Mixed Model (lme)
- Dependent Variable: log(TrialDuration)
- Fixed Effects: PrePostControlCondition
- Random Effects: $\sim 1 +$ Participant | Participant
- Model Fit Statistics: AIC = -349.665, BIC = -327.0927, logLik = 180.8325

Results:

- Intercept: Estimate = 3.490358, SE = 0.10280603, t-value = 33.95090, $p - value <$ 0.001
- PrePostControlCondition: Estimate = -0.041963, SE = 0.01421692, t-value = -2.95161, $p - value = 0.0034$

Effects:

- Participant (Intercept): StdDev = 2.893858e-01
- Participant: StdDev (Intercept) correlation = 0
- Residual: StdDev = 1.271600e-01

Data Samples:

- Number of Observations: 320
- Number of Groups: 8

**Effect of training phase (pre/post training condition) on number of collisions**

Model Specification:

- Model: Negative Binomial Regression (glm.nb)
- Dependent Variable: Collisions
- Fixed Effects: PrePostTrainingCondition
- Random Effects: $\sim 1 +$ ParticipantID | ParticipantID

- Model Fit Statistics: Null Deviance: 291.34 on 319 degrees of freedom, Residual Deviance: 278.05 on 318 degrees of freedom, AIC: 762.19

Results:

- Intercept: Estimate = 0.02469, Std. Error = 0.12360, z value = 0.200, $p-value =$ 0.841657
- PrePostTrainingCondition: Estimate = -0.69315, Std. Error = 0.19145, z value = -3.621, $p-value = 0.000294$
- Random Effects: $\sim 1 + Participant \mid Participant$ (Not defined due to singularities)

Additional Information:

- Theta: Estimate = 0.681, Std. Error = 0.147
- Number of Fisher Scoring iterations: 1
- 2 x log-likelihood: -756.191

No estimates can be given for the random effect for this model. This is likely caused due to the number of subjects being too small.

**Effect of control phase (pre/post control condition) on number of collisions**

Model Specification:

- Model: Negative Binomial Regression (glm.nb)
- Dependent Variable: Collisions
- Fixed Effects: PrePostControlCondition
- Random Effects: $\sim 1 +$ Participant | Participant
- Model Fit Statistics: Null Deviance: 251.63 on 319 degrees of freedom, Residual Deviance: 251.37 on 318 degrees of freedom, AIC: 749.11

Results:

- Intercept: Estimate = -0.2469, Std. Error = 0.1500, z value = -1.645, $p-value =$ 0.0999
- PrePostControlCondition: Estimate = -0.1098, Std. Error = 0.2144, z value = -0.512, $p-value = 0.6085$
- Random Effects: $\sim 1 +$ ParticipantID | ParticipantID

Additional Information:

- Theta: Estimate = 0.4307, Std. Error = 0.0814
- Number of Fisher Scoring iterations: 1
- 2 x log-likelihood: -743.1110

No estimates can be given for the random effect for this model. This is likely caused due to the number of subjects being too small.

**Effect of training phase (pre/post training condition) on head-centric DFoV**

Model Specification:

- Model: Linear Mixed Model (lme)
- Dependent Variable: HeadCentricDFoV
- Fixed Effects: PrePostTrainingCondition
- Random Effects: $\sim 1 +$ Participant | Participant
- Model Fit Statistics: AIC = -345.4398, BIC = -324.6062, logLik = 178.7199

Results:

- Intercept: Estimate = 1.0161667, Std.Error = 0.01021678, DF = 231, t-value = 99.46056, $p-value < 0.001$
- PrePostTrainingCondition: Estimate = -0.0196667, Std.Error = 0.01444871, DF = 231, t-value = -1.36114, $p-value = 0.1748$

Effects:

- participant (Intercept): StdDev = 1.301521e-06
- participant: StdDev (Intercept) correlation = 0
- Residual: StdDev = 1.119192e-01

Data Samples:

- Number of Observations: 240
- Number of Groups: 8

**Effect of control phase (pre/post training condition) on head-centric DFoV**

Model Specification:

- Model: Linear Mixed Model (lme)
- Dependent Variable: HeadCentricDFoV
- Fixed Effects: PrePostControlCondition
- Random Effects: $\sim 1 +$ Participant | Participant
- Model Fit Statistics: AIC = -225.9001, BIC = -206.1403, logLik = 118.95

Results:

- Intercept: Estimate = 1.0163366, Std.Error = 0.01294036, DF = 193, t-value = 78.54007, $p - value < 0.001$
- PrePostControlCondition: Estimate = -0.0160366, Std.Error = 0.01834613, DF = 193, t-value = -0.87412, $p - value = 0.3831$

Effects:

- Participant (Intercept): StdDev = 1.442753e-06
- Participant: StdDev (Intercept) correlation = 0
- Residual: StdDev = 1.300490e-01

Data Samples:

- Number of Observations: 201
- Number of Groups: 7

**Effect of training phase (pre/post training condition) on world-centric DFoV**

Model Specification:

- Model: Linear Mixed Model (lme)
- Dependent Variable: WorldCentricDFoV
- Fixed Effects: PrePostTrainingCondition
- Random Effects: $\sim 1 +$ Participant | Participant
- Model Fit Statistics: AIC = -221.0613, BIC = -200.2277, logLik = 116.5307

Results:

- Intercept: Estimate = 0.97800, Std.Error = 0.01326768, DF = 231, t-value = 73.71298, $p - value < 0.001$
- PrePostTrainingCondition: Estimate = 0.06625, Std.Error = 0.01876333, DF = 231, t-value = 3.53082, $p - value = 0.0005$

Effects:

- Participant (Intercept): StdDev = $2.303544e - 06$
- Participant: StdDev (Intercept) correlation = 0
- Residual: StdDev = 0.1453401

Data Samples:

- Number of Observations: 240
- Number of Groups: 8

**Effect of control phase (pre/post control condition) on world-centric DFoV**

Model Specification:

- Model: Linear Mixed Model (lme)
- Dependent Variable: WorldCentricDFoV
- Fixed Effects: PrePostControlCondition
- Random Effects: $\sim 1 +$ Participant | Participant
- Model Fit Statistics: AIC = -90.34245, BIC = -70.58262, logLik = 51.17122

Results:

- Intercept: Estimate = 0.9865347, Std.Error = 0.01819139, DF = 193, t-value = 54.23087, $p - value < 0.001$
- PrePostControlCondition: Estimate = 0.0448653, Std.Error = 0.02579074, DF = 193, t-value = 1.73959, $p - value = 0.0835$

Effects:

- Participant (Intercept): StdDev = $4.234727e - 06$
- Participant: StdDev (Intercept) correlation = 0
- Residual: StdDev = 0.1828212

Data Samples:

- Number of Observations: 201
- Number of Groups: 7

##### QQ-plots

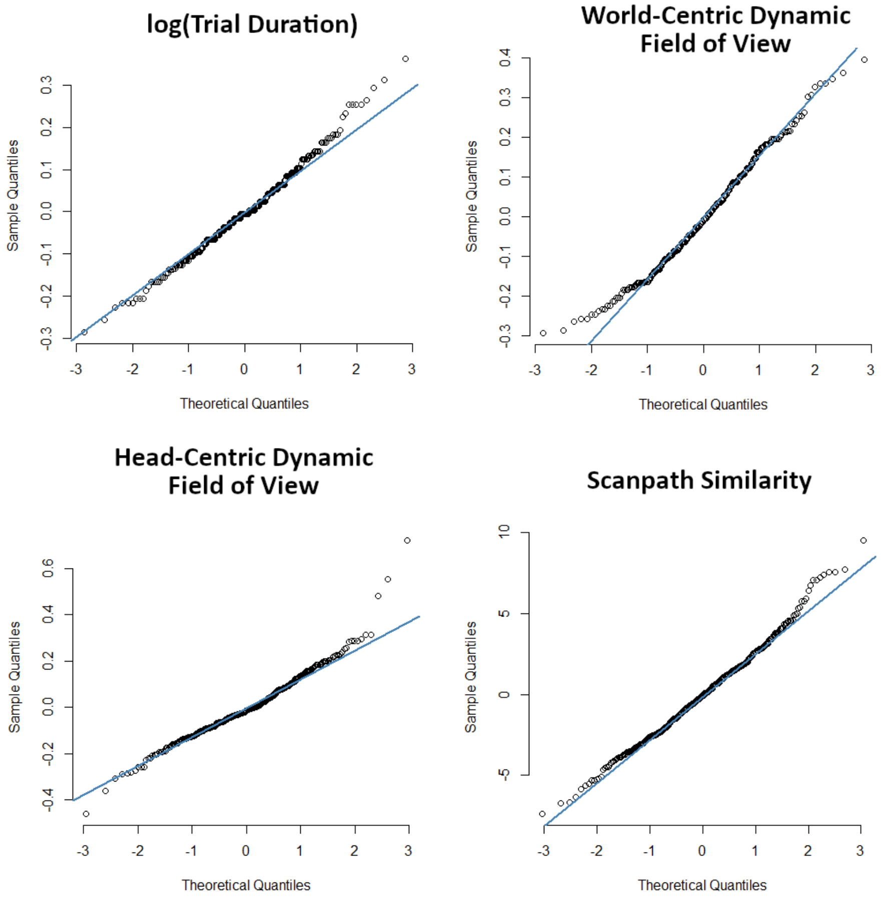


Figure 14: QQ-plots of the residuals of different result parameters of the real-world obstacle course.
